# LLM-AIx: An open source pipeline for Information Extraction from unstructured medical text based on privacy preserving Large Language Models

**DOI:** 10.1101/2024.09.02.24312917

**Authors:** Isabella Catharina Wiest, Fabian Wolf, Marie-Elisabeth Leßmann, Marko van Treeck, Dyke Ferber, Jiefu Zhu, Heiko Boehme, Keno K. Bressem, Hannes Ulrich, Matthias P. Ebert, Jakob Nikolas Kather

## Abstract

In clinical science and practice, text data, such as clinical letters or procedure reports, is stored in an unstructured way. This type of data is not a quantifiable resource for any kind of quantitative investigations and any manual review or structured information retrieval is time-consuming and costly. The capabilities of Large Language Models (LLMs) mark a paradigm shift in natural language processing and offer new possibilities for structured Information Extraction (IE) from medical free text. This protocol describes a workflow for LLM based information extraction (LLM-AIx), enabling extraction of predefined entities from unstructured text using privacy preserving LLMs. By converting unstructured clinical text into structured data, LLM-AIx addresses a critical barrier in clinical research and practice, where the efficient extraction of information is essential for improving clinical decision-making, enhancing patient outcomes, and facilitating large-scale data analysis.

The protocol consists of four main processing steps: 1) Problem definition and data preparation, 2) data preprocessing, 3) LLM-based IE and 4) output evaluation. LLM-AIx allows integration on local hospital hardware without the need of transferring any patient data to external servers. As example tasks, we applied LLM-AIx for the anonymization of fictitious clinical letters from patients with pulmonary embolism. Additionally, we extracted symptoms and laterality of the pulmonary embolism of these fictitious letters. We demonstrate troubleshooting for potential problems within the pipeline with an IE on a real-world dataset, 100 pathology reports from the Cancer Genome Atlas Program (TCGA), for TNM stage extraction. LLM-AIx can be executed without any programming knowledge via an easy-to-use interface and in no more than a few minutes or hours, depending on the LLM model selected.

## Introduction

### Development of the protocol

Medical free text contains essential information, such as details about patient characteristics and therapy course and maps the patient journey substantially better than structured medical information from electronic health records alone ^1–3^.This medical free text contains the main reasoning as well as observations from medical staff within a variety of different report types, such as clinical letters as well as documentation of different diagnostic and therapeutic procedures ^4^. In its unstructured form, text is not available for quantitative analysis and is therefore not accessible for research, quality analysis or interoperable data exchange ^5^. Forcing medical staff into structured documentation, however, is not feasible due to time constraints and shortage of personnel in the healthcare system. This leads to an increasing documentation burden and decreases the time available for actual patient care ^6^. Therefore, systematically extracting information from free text is crucial for the medical field: It enables researchers to investigate rare diseases ^7^, allows better tracking, overview, and exchange of patient information among different inpatient and outpatient providers via a comprehensive health record, and systematic quality control assessment ^8,9^ .

Previous methods to mine medical free text fall short because they are either not capable of processing large amounts of text and have limited capabilities to grasp context, or need task-specific fine-tuning ^10^, whereas our method solely relies on in-context learning of large language models. In-context-learning enhances LLMs’ performance on new tasks by using examples or step-by-step instructions within the prompt ^11,12^. Additionally, narrative medical text comes from various source systems, which complicates a streamlined processing. Some reports may only be accessible in portable document format (PDF) from the clinical information system (CIS), others originate from secondary software in a variety of different formats.^13^ Data transformation processes to harmonize all the data formats from their source systems within one central database are not ubiquitously established ^14^. We therefore present an open-source, LLM-based pipeline which tackles these challenges in medical information extraction (IE). Additionally, our pipeline extracts structured information elements that can be flexibly defined by the user. This is advantageous compared to traditional IE, where predefined categories and relationships are extracted. Our approach offers a highly flexible process for handling large-scale unstructured data ^15^.

Our pipeline is able to transform various types of unstructured medical text data—such as clinical notes, procedure reports or entire clinical letters—into structured CSV format, suitable for quantitative analysis. This development was motivated by the need for a scalable solution that accommodates the technical expertise and deep medical domain understanding required for effective data utilization in healthcare. The method has been developed, applied and tested for several use cases, namely extracting **suicidality** of psychiatric admission notes ^16^, tested with different Large Language Models from Meta AI (Llama-2 models). Additionally, we extracted several **symptoms and diagnoses** for detection of decompensated liver cirrhosis from emergency room (ER) admission notes ^17^. Furthermore, we applied the pipeline for extracting **adverse events** from endoscopy reports of endoscopic mucosal resection (EMR) and colonoscopies ^18^. All of these proof-of-concept studies led to the development of the entire pipeline presented here, which comprise an intuitive graphical user interface (GUI), data preprocessing, LLM-based IE as well as automated evaluation of the process within one pipeline. Previously, we introduced the LLM Anonymizer, which is a special case for IE with the purpose of **anonymizing** medical reports ^19^.

The latest open-source LLMs can easily be implemented within the pipeline, which also facilitates benchmarking of different models in accurately extracting relevant entities and information based on the specific needs of requestors. Currently, all models available in Generative Pre-Trained Transformers (GPT)-Generated Unified Format (GGUF) can be included in the pipeline, such as Llama-2 with 7 billion parameters (7b), Llama-2 70b, Llama-3 8b, Llama-3 70b, Llama-2 “Sauerkraut” 70b, Phi-3, Mistral 7b and many more. By producing outputs in a CSV format, we enable seamless integration with existing data analysis tools and workflows, facilitating quantitative analysis without the need for specialized computational skills. As an example, existing databases such as cancer registries or clinical databases could be filled with the help of our pipeline.

### Overview of the protocol

The protocol consists of four main stages: 1) Problem definition and data preparation 2) Data preprocessing, 3) LLM-based IE and 4) Output evaluation (**Figure 1**). The protocol facilitates any kind of IE from medical free text documents, with a variety of input formats possible. It is easy to use for clinical researchers without NLP expertise and allows the application of the latest LLMs for medical IE. We have shown that the protocol is broadly applicable to any kind of medical text data. The protocol is available in an open-source codebase on github (available at https://github.com/KatherLab/LLMAIx). Additionally, our method can be implemented on low hardware resources (e.g., a single graphical processing unit (GPU) with 48GB video random-access memory (VRAM)), making it more accessible and cost-effective compared to systems with higher computational demands.

**Figure 1.**
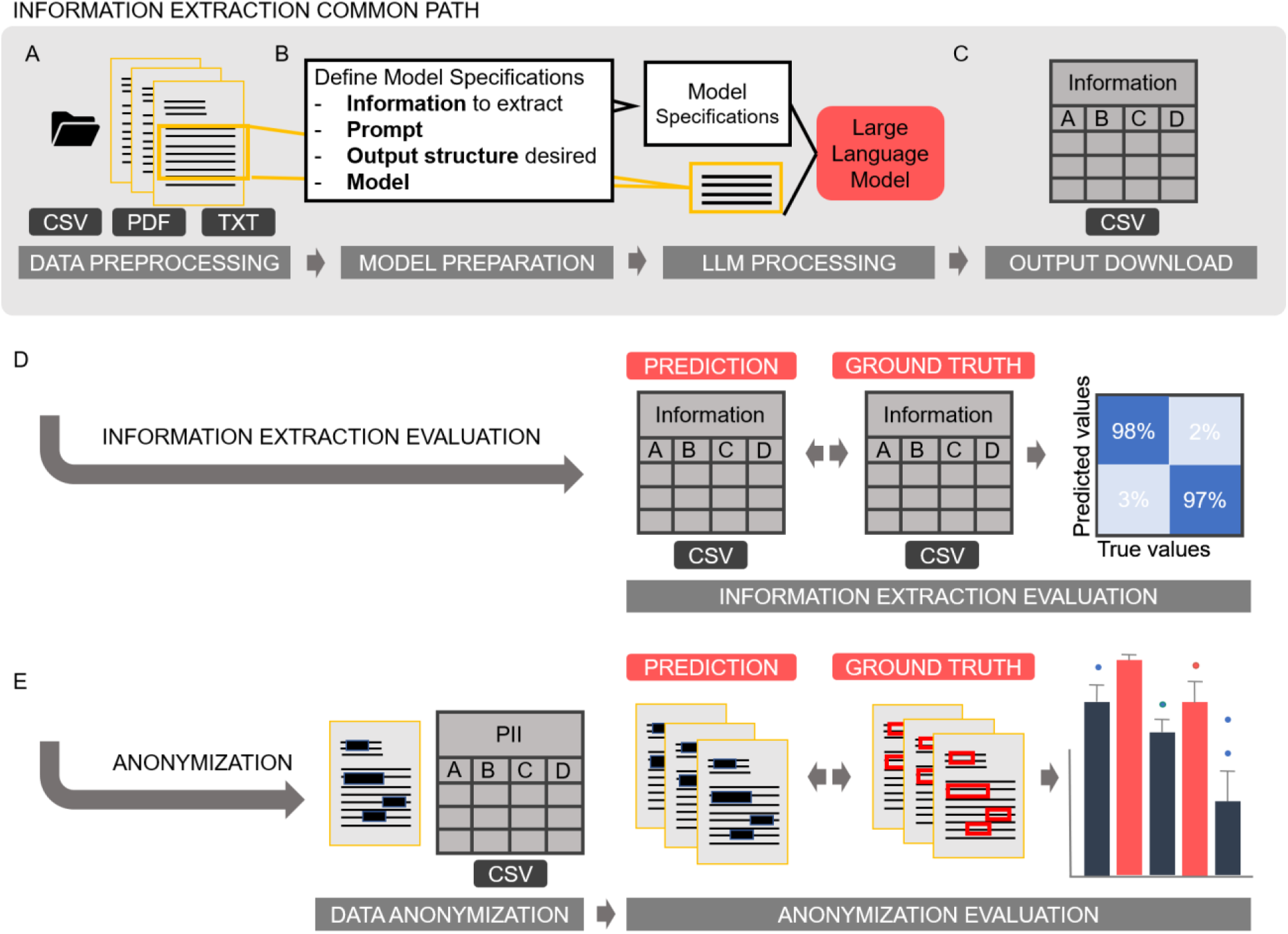
Information Extraction Workflow. **A** The information extraction pipeline follows a common path that includes data preprocessing, optional Optical Character Recognition (OCR), document splitting, and support for various file formats (CSV, Excel, PDF, or TXT). **B** After pre-processing, users can specify model parameters such as hyperparameters, prompts, and desired output structure. Once these are defined, the LLM-based information extraction process begins. **B** The resulting ZIP file contains the output CSV with LLM predictions of the desired information and the original reports. The evaluation process offers two options. **D** If the pipeline is used for information extraction, it identifies and extracts the required information into a CSV file. This extracted CSV file can then be compared to a ground truth CSV file. Confusion matrices and comprehensive performance metrics are generated to visualize and evaluate the pipeline’s performance. **E** If the pipeline is used for document anonymization, the original documents are redacted to obscure personal identifiers and can be compared to annotated data files. The pipeline automatically generates confusion matrices that visualize matching and mismatching characters, facilitating easy performance evaluation. The anonymization part of this figure is based on the workflow depiction of our previous publication^19^.

### Applications of the method

The pipeline has been applied in several clinical use cases to demonstrate its versatility and effectiveness, and can be applied for any use case where quantifiable, structured data is required from unstructured medical text. Unlike traditional NLP methods that often require specific training and fine-tuning ^20^, this pipeline utilizes LLMs which excel in zero-shot applications, which refers to the ability of LLMs to make predictions on data that was not encountered during training, without requiring any task-specific fine-tuning. This makes LLMs ideal for processing a wide range of medical documents.

Furthermore, the performance of LLMs can be optimized through direct interaction via prompting and prompt engineering ^21,22^. This facilitates in-context learning, a methodology wherein the user presents the desired output to the model and provides one or multiple examples of the correct solution ^11^. This approach is entirely based on model inference, eliminating the need for extensive retraining of the model.

The procedure can be applied for a variety of purposes:

1. Interdisciplinary collaboration. When exchanging healthcare data across multiple centers for research, a uniform data standard is key. LLM information extraction based on a predefined data standard could support interdisciplinary cooperation without the need of exchanging original text data, which may contain sensitive information that may remain on the sites where the data emerges.
2. Clinical research. Quantitative research is only possible with quantitative data and cannot be performed with quantitative information hidden in free text data. Current practice is manual extraction of information from medical text by medical documentalists or scientific assistants. This is, however, time consuming and is complicated by personnel shortage in the healthcare sector. Additionally, the tool could support filling patient registries such as cancer registries and clinical trial documentation (e.g. by structured extraction of adverse events from free text clinical notes).
3. To build quantitative downstream models. For example, to predict certain outcomes from other data modalities, such as radiology images, one needs outcome information about the respective patients which is usually hidden in free-text documentation. This information can be extracted with our pipeline and then serve as a label for predictive machine learning model training ^23–25^.
4. Quality assurance and auditing. The tool could help to complete clinical data used for measuring quality of care.
5. EHR Data integration. The structured information could enrich patients’ electronic health records to then reflect a more complex picture of patient history and treatment within an interoperable and accessible way for all healthcare providers.

### Comparison with other methods

Until now, due to the shortcomings of other methods, the gold standard in medical IE is labor intensive, manual IE by medical documentaries or medical staff. Traditional methods such as machine learning named entity recognition (NER) methods typically require the extraction of fixed entities and offer limited flexibility. To name an example, they extract all names, dates and locations from a text, without being able to interpret the context. If only the surgery date was needed from a report, this could not easily be identified among all dates mentioned within a text. In contrast, our LLM-based approach allows for the flexible definition of entities to be extracted through advanced prompt engineering and in-context learning capabilities. This adaptability makes it more suitable for the dynamic and varied needs of medical data analysis.

### NLP in the pre-transformer area - Beginnings of pre-trained models

Initially, IE relied on hand-crafted rules and required extensive manual efforts to define patterns which were limited in their adaptability to different domains ^26,27^. Machine learning techniques, which used labeled data to train models, improved the IE performance in the NLP domain ^28^. These methods leveraged features as part-of-speech tags; such as nouns, verb or adjectives ^29^, syntactic structures; such as noun phrases, verb phrases and adjective phrases ^30^, and lexical cues, which are words indicating a specific relation and entity, such as intensifiers like “very” in the sentence “she was very happy” ^31^, to improve the accuracy of entity recognition and relation extraction. Notable algorithms like Hidden Markov Models (HMMs) and Conditional Random Fields (CRFs) were widely adopted for these tasks ^32,33^. Non-neural methods such as n-gram models ^34–36^, and neural network methods, particularly recurrent neural networks (RNNs) and convolutional neural networks (CNNs) were able to improve capturing contextual information ^37^. However, labeled data is scarce, particularly in the medical domain, and as a result, unsupervised and semi-supervised learning approaches were advanced simultaneously. They aim to utilize large amounts of unlabeled text to automatically discover patterns in the data. Word embeddings, which are representations of words in continuous vector spaces, such as Word2Vec or GloVe, further enhanced the generalizability across different contexts ^38^. ULMFit followed as one of the first approaches for pre-trained models ^39^. However, all of these methods suffered from limited context understanding in document level, do not capture polysemy, have a fixed vocabulary size, require large text corpora for training and were so far insufficient for IE in the medical field ^40^.

### New prospects with LLMs

The development of the transformer architecture, a deep learning architecture that is based on multi-head attention ^41,42^, substantially changed the NLP landscape: Especially the introduction of Bidirectional Encoder Representations from Transformers (BERT) and Generative Pre-trained Transformer (GPT) advanced the field. These models capture language patterns and context, and subsequent fine-tuning of pre-trained models on specific tasks achieved good results in entity recognition ^43^. BERT-based models have also been established for the biomedical domain (BioBERT, SciBERT, ClinicalBERT, BioMedRoBERTa) and tested on several benchmark datasets (GLUE, MuliNLI, SQuAD) ^44,45^. Nevertheless, BERT models require fine-tuning for successful IE, which requires procedure and programming knowledge and have very limited context length ^10^.

LLMs, which have larger parameter sizes than BERT-based language models, have shown great potential in classical IE tasks ^46^. They offer a high zero-shot performance and shift the task solving field towards immediate prompt engineering instead of fine-tuning and model training. In-context-learning, which does not alter the model’s weights and has the advantage of using purely natural language, potentially allows medical staff without programming knowledge to seamlessly integrate these tools into their daily routines. Furthermore, it provides maximum flexibility to extract contextually relevant information as specified by the requester, requiring minimal programming knowledge, making it ideal for the information extraction process in the medical field ^47^. The strength of our approach lies in its robust performance across datasets of any size, ensuring efficiency and accuracy whether analyzing a single report or aggregating insights from a vast collection of documents.

### Experimental design

To validate the efficacy of our protocol, we conducted experiments across different datasets in different languages and clinical settings. Each use case was designed to test the protocol’s ability to accurately and efficiently process unstructured text into structured data while addressing specific clinical questions. The performance metrics included accuracy, sensitivity, specificity, F1-score and precision, and the ability to maintain data integrity and privacy.

### Expertise needed to implement the protocol

We have designed the pipeline to require almost no programming knowledge with a user interface that allows intuitive data processing for non-technical users, however, pipeline setup requires some knowledge about virtual environments and navigating a terminal. Additionally, a useful application requires domain knowledge, therefore it is crucial that medical experts clearly define the entities of interest to enable concise and effective prompting, which is central to the protocol’s operation. This requirement highlights the importance of having a well-understood and agreed-upon definition of the entities among the clinical team members to facilitate the accurate extraction of information.

### Limitations

While our pipeline significantly enhances the accessibility and utility of unstructured medical text data, it does have limitations:

- Dependence on High-Quality Data Inputs: The effectiveness of the LLM is contingent on the quality and diversity of the input data. Handwritten documents and poorly scanned files may not be effectively processed by the implemented Optical Character Recognition (OCR) engines.
- Computational Resources: Despite the possibility to run the pipeline on consumer hardware, the necessity for a GPU with substantial VRAM may limit implementation in re-source-constrained settings. Models with larger parameter sizes (such as Llama 3.1 405B) may require additional hardware to be tested.
- LLM inherent constraints: LLMs may generate information or statements that are factually incorrect, misleading, or fabricated, so called “hallucinations”. They can be mitigated by adjusting hyperparameters and providing proper in-context learning, though it cannot be completely eliminated. However, LLM-AIx can still reduce IE time, even with a human in the loop. When using a homogeneous dataset, the error rate in IE can be considered comparable to that of a small evaluation subset ^18^.

## Materials

### Data

The unstructured text data used to run the pipeline can have different formats. Our pipeline allows processing of portable document format (PDF), raw text (TXT) or comma-separated values (CSV) as well as EXCEL files.

### Hardware

The pipeline can be run fully locally on consumer hardware (such as NVIDIA RTX 4090 or Apple Silicon M2 of a macbook). We ran the pipeline with one Graphics Processing Unit (GPU), equipped with 48 gigabytes (GB) of video random-access memory (VRAM) with a NVIDIA RTX A6000. In theory, the pipeline can be deployed on consumer hardware with any GPU (minimum 12 GB of VRAM). Processing times, context length and memory may limit the deployment on consumer hardware to smaller LLMs and datasets.

To enable use on comparatively low-resource hardware, we employed only quantized models (4- and 5-bit quantization), which are smaller than unquantized LLMs but maintain comparable performance ^48^.

### Software

The pipeline can be used through a graphical user interface without any programming knowledge. It can be downloaded as a Docker image for a quick setup, including all its dependencies except the model files in GGUF format.

Alternatively, manual setup is possible by installing the required python packages as well as additional software packages (tesseract, llama.cpp) as it is described in the README.md file (https://github.com/KatherLab/LLMAIx). All software packages require a minimum of Python 3.12.

The data preprocessing stage includes different options of OCR for processing image-only PDFs. We implemented the popular open source OCR “tesseract” via the package OCRmyPDF, as well as potentially superior alternatives such as “surya” ^49^, which can be selected by the User. Default OCR is tesseract.

The protocol adopts the llama.cpp framework which enables the application of a variety of LLMs, formatted in GGUF. It allows LLM inference with state-of-the-art performance on a variety of hardware locally and in the cloud ^50^. It is an open-source project that enables the use of Llama (Large Language Model Meta AI) models in C++.

### Equipment Setup

To set up the pipeline, two main steps are necessary: 1) Model download, 2A) Docker pipeline setup or 2B) Manual pipeline setup:

1) Download the desired models in GGUF format onto your local system.

2A) Edit docker-compose.yml with the correct model path and follow the instructions. Then run the docker image as described in the README.md.

2B) Download the pre-built llama.cpp from Github and follow the installation instructions ^50^. Then clone the pipeline-Github repository. Create a virtual environment and install all necessary python packages within the environment. The detailed implementation of all dependencies and setup is described in the README.md file.

## Procedure

### Stage 1: Problem definition and data preparations

⚫ TIMING

🔹 TROUBLESHOOTING

⚑PAUSE POINT

1. ***Define the use case:***

- TIMING: variable Preparation for utilizing our protocol involves users to define their specific extraction tasks clearly. This includes specifying the nature of the information to be extracted (for example, identifying complications from endoscopy reports), the format and volume of the data under analysis, and the desired output categories for subsequent analysis. To accommodate documents in various formats (TXT, PDF, or CSV), our protocol standardizes the data into a uniform format (CSV) through automatic conversion and compilation. This standardization is critical for ensuring consistent analysis across diverse datasets.
2. ***Assess the input data:***

- TIMING: variable Identify the raw text data and the format that is available. A patient can have multiple documents, the processing happens per document. If the user plans to process CSV or EXCEL files, the text needs to be filled in a dedicated column, called “report”. Additionally, each text document needs a unique ID in the column “id”. If the user plans to upload single files for each document (e.g. PDF), the files need to be named by the unique ID. Consistency in data labeling needs to be ensured in this data preparation step.
3. ***Define the validation strategy:***

- TIMING: variable This can either be done with document-wide labels (IE-Pipeline) or annotated text data (IE-Anonymizer).
4. ***Prepare ground truth data:***

- TIMING: variable If automated evaluation of the LLM IE pipeline will be performed, a ground truth table of the data or a data subset needs to be provided. If there are unique labels per document, the ground truth needs to be provided in tabular format (CSV or EXCEL), containing all variables that should be extracted as columns and the corresponding ground truth, with the same characteristics as defined for IE. If text annotations and respective labels are supposed to be compared to the LLM output, annotated JSON files (one for each document) need to be zipped. We recommend annotation with INCEpTION ^51^, for which the annotation comparison of this pipeline has been optimized.
5. ***Download the desired LLMs***

- TIMING: variable If not performed beforehands, the user needs to create a folder on the local computer and download the desired LLMs to that folder. All models must be downloaded in GGUF format and can be accessed via huggingface.com. Hugging Face is a company and open-source community that provides a wide range of tools and libraries for NLP and machine learning. We tested several models within the pipeline, among them Llama-3.1 ^52^ and Llama-3 ^53^ models, Mistral ^54^, Llama-2 ^55,56^, Gemma ^57^ and Phi-models ^58^.
6. ***Setup for running the pipeline***

- TIMING: ∼5 minutes The pipeline can either be run via docker image or set up and started manually. Detailed descriptions can be found in the README.md. The procedure will be described with the Docker version. Docker needs to be installed ^59^.

### Stage 2: Data preprocessing

7. ***Pipeline initiation:***

- TIMING: ∼1 min ***A Docker*** The docker-compose.yml must be adapted with the correct model path as described in the README.md. Then, run the docker image with “docker-compose up”. ***B Manually*** To initiate the pipeline, the user activates their virtual environment and navigates into the repository via terminal. There, the app can be started with the terminal command “python app.py –model_path USERMODELPATH”. The application will then be loaded on the local server and can be run browser based. Click on the link provided in the terminal. (Default: http://localhost:5000). The user chooses the IE-mode based on preferences, either IE or anonymization mode can be chosen **(Figure 2)**.
8. ***Preprocess the data:***

- TIMING: variable depending on the amount of reports which necessitate OCR recognition. After the data has been prepared, it can be preprocessed. The “Preprocessing” tab allows to upload the data files by clicking the “Upload Data” button. PDF files can be uploaded, both “image-only” or PDF documents with a text layer. The name of the PDFs should contain the document id and match the annotated ground truth file id. Additionally, EXCEL or CSV files can be uploaded. They also need to contain an “id” column and a “report” column, containing one report per line. Once all documents are uploaded, the user can select the desired OCR method from a drop-down menu and specify the number of characters. If needed, the document will then be split to accommodate the limited context windows of certain models. After clicking the “Preprocess Files” button, a progress bar appears and indicates the preprocessing status. As soon as finished, the preprocessed data needs to be downloaded as a zipped folder. This zip folder contains all original documents in a separate PDF file as well as a CSV file. **(Figure 2)** ⚑🔹

**Figure 2.**
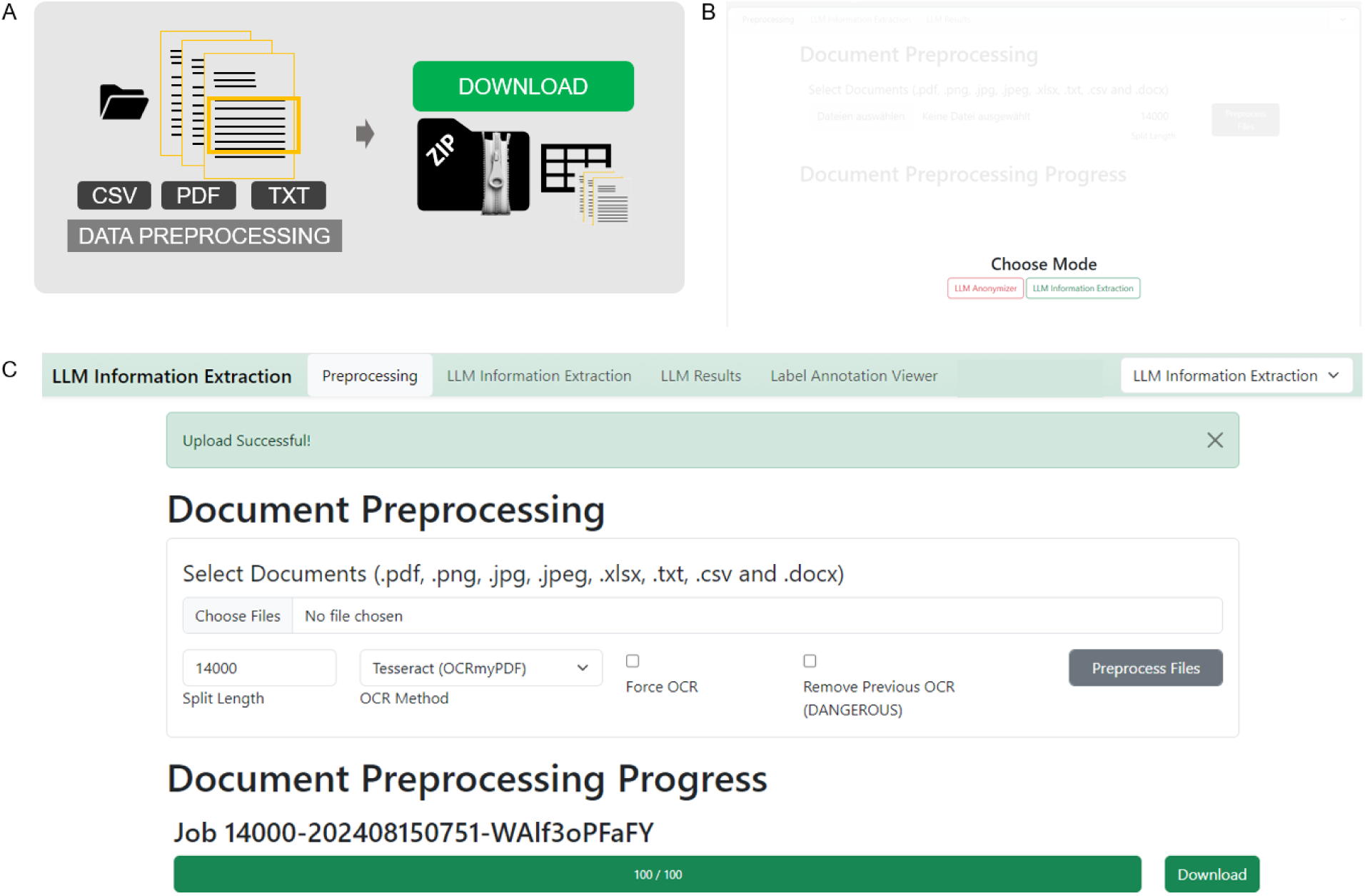
Preprocessing Procedure GUI - Stage 2, 7-8. **A** Schematic depiction of the preprocessing stage. Files can be uploaded and will be split to smaller document chunks if necessary according to the split size determined. A zip file containing original documents and a CSV file that organizes all documents for LLM processing can be downloaded in the end **B** When the browser based application is started, the mode of action can be chosen, which is either the “LLM Anonymizer” or the “LLM Information Extraction”. Both modes have the same preprocessing and data processing, however the evaluation part differs. Modes can be switched at any time during the process by adapting the drop-down menu in the upper right corner. **C** The user’s documents can be uploaded in the preprocessing step. Images, as well as portable document format (PDF), raw text, as well as excel or comma separated value (CSV) files are allowed. The split size, which can be determined according to the expected context window size of the LLM, is 14000 characters per default but can be adapted if necessary. By clicking the “Preprocess Files” button, the preprocessing starts. Progress is indicated with a progress bar. The “Download” button allows you to download and store the preprocessed zip file which will be needed in further processing steps.

### Stage 3: LLM-based information extraction

9. Prepare the LLM based information extraction *(Figure 3)*

- TIMING: Depends on the amount of variable and complexity of the prompt, ∼2 min. Model Selection The user specifies the desired model by choosing it from the drop-down menu. We have predefined the most common open source models. Additional models can be added by downloading them to the predefined model folder and adding them to the “yaml file”. Upload the preprocessed zip file With the “upload” button, the preprocessed zip file can be uploaded for further processing. Prompt definition The prompt can be defined in the “Prompt” field and can be customized according to the user’s needs. At the location of “report” in parenthesis, the respective report will be inserted, therefore this element needs to stay within the prompt as is. The location can be varied upon necessity. We recommend a prompt that is structured in two main parts: Giving background to the model and instructing the model with the task. If the IE-task is more complex, it can be helpful to insert examples for few-shot prompting that leverage in-context learning of the models. It is important to demonstrate examples in the same format as the output is desired. Grammar specification The section “Grammar” contains a JSON schema defining the output structure of the model. The grammar based sampling approach that is applied here enables the user to flexibly define the features to be extracted from the text and their possible outcomes. The desired model output JSON schema can either be adapted manually (error-prone and therefore not recommended) or with the “Grammar Builder”. The user can assign a label name for the information to be extracted and select the desired output format (string, boolean, categories, number) and further specify the categories (in a comma-separated list) and the character length of the string and number. Once defined, the grammar can be downloaded and stored as a CSV file by clicking the “Save Configuration” button. Whenever the same configuration is required again, it can be uploaded via the “Load Configuration” button. Once the grammar is set up correctly, the user loads it into the “Grammar” section with the button “Generate Full Grammar”. If prompt, grammar and hyperparameters are correctly defined and the preprocessed file is uploaded, clicking the button “Run LLM Processing” initiates the process.
10. Run the LLM based information extraction

- TIMING: variable, depending on model size and number of reports. First, the respective model will be uploaded onto the local server. This might take a few seconds up to several minutes, depending on the model size and hardware specifications. When the model is loaded, processing starts and a progress bar indicates the remaining amount of reports and time for the process. The remaining time is corrected after each report is processed. After the process is finished, the user can download and store the processed zip file. This file contains all original reports as PDFs, the preprocessed CSV as well as an output CSV that contains all LLM answers as well as meta-information including prompt and hyperparameter settings. ⚑🔹

**Figure 3.**
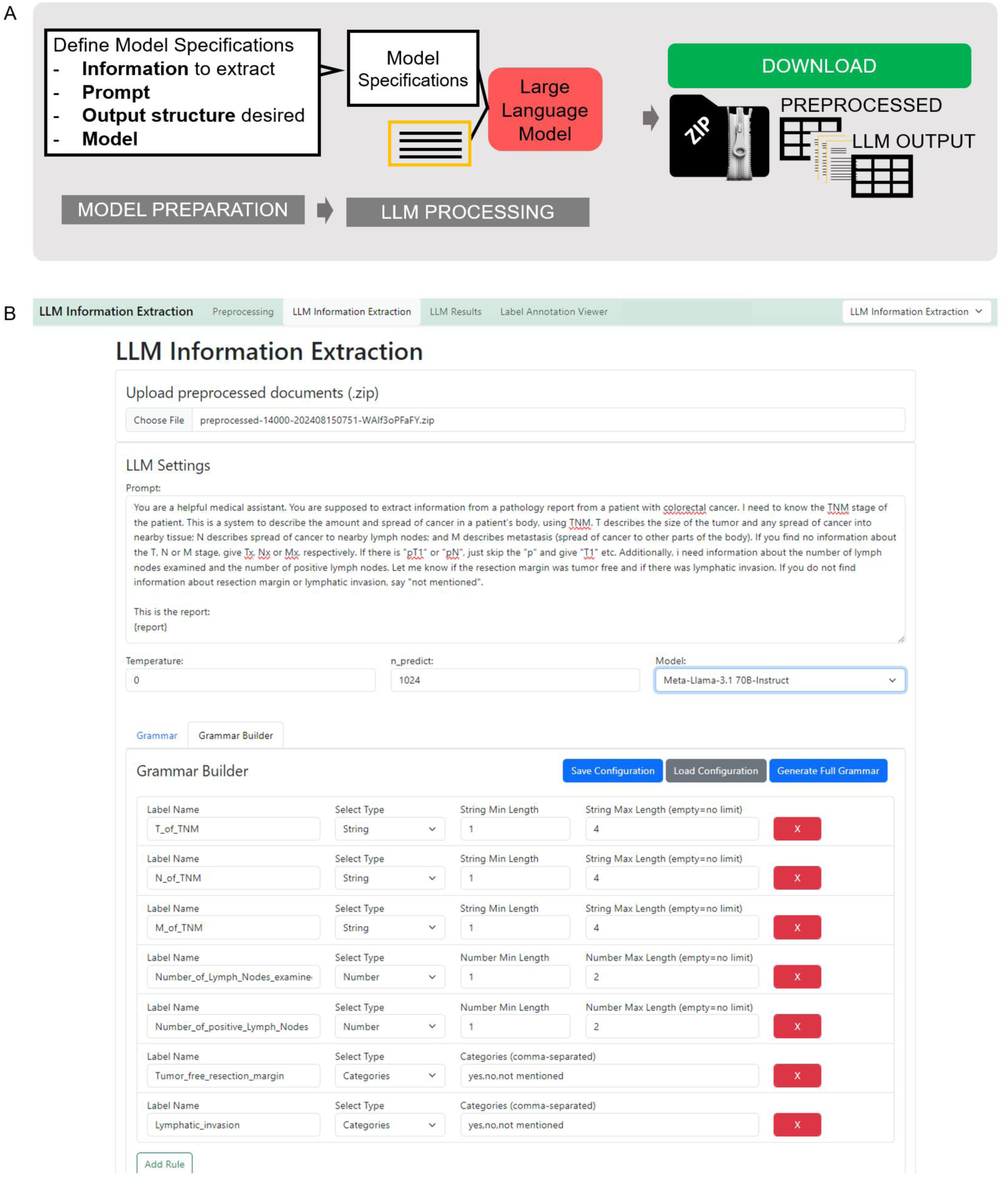
LLM-processing Procedure - Stage 2, 7-8. **A** Schematic depiction of the LLM-processing stage. The large language model (LLM) based information extraction requires uploading of the preprocessed zip file. Then LLM Settings can be determined. The prompt field allows inserting a specific prompt. At the “{report}” indicator, your original report text will be inserted. Additionally, LLM hyperparameters can be set (Temperature and tokens to be predicted (n_predict)) and the desired model can be chosen via a dropdown menu. To ensure a consistent output structure, the model can be given a JavaScript Object Notation (JSON) schema. This can either be defined manually, which is prone to errors. Therefore, we implemented a “Grammar Builder” that allows to define your feature name and values to be extracted. The grammar configuration shown in **B** can be downloaded and stored on your local computer and loaded whenever needed. With the button “Generate Full Grammar”, it will be loaded to the processing mask. Afterwards, “Run LLM Processing” starts the information extraction. The model chosen will then be loaded on the local Graphical Processing Unit (GPU), before the information extraction starts. This is indicated by a loading circle in the Graphical User Interface (GUI) or the uploading dot indicators in the terminal. As soon as the model is successfully loaded to the GPU, the LLM-based processing starts and progress is indicated with a progress bar.

### Stage 4: Output evaluation

⚫ TIMING

🔹 TROUBLESHOOTING

11. ***Run evaluations***

- TIMING: variable, depending on number of reports. ∼1-2 sec/report ***A Run evaluation in Information Extraction Mode*** The “Label Annotation Viewer” enables the uploading of both the output ZIP file and the ground truth CSV file. It is essential to ensure that the IDs and variable names are unique and matching in both files. (**Figure 4 A**)The process can be initiated by clicking “Label Annotation Metrics Summary” to obtain all metrics and “Label Annotation Viewer” to review results on a document-by-document basis. Before results are obtained, data types of the extracted variables have to be confirmed to ensure proper evaluation (**Figure 4 B**). ***B Run evaluation in Anonymizer Mode*** To evaluate the output in the Anonymizer mode, the output zip file can equally be uploaded. The ground truth needs to contain annotations for the individual reports in JSON format. For annotation tasks, we used the open source annotation tool “INCEpTION” ^51^. The annotated JSON exports need to be zipped and can then be uploaded equally to the IE evaluation. 🔹
12. ***Revise Metrics and Files***

- TIMING: variable, up to the user’s preferences The “Label Annotation Summary” provides global metrics for the experiment by comparing the output data to the ground truth data. For boolean IE, it presents a comprehensive set of metrics, including Accuracy, F1 Score, Precision, Recall, False Positive Rate, and False Negative Rate, both across all reports and variables, and for each variable individually. A confusion matrix visualizes true and false positives and negatives (**Figure 5A**). For categorical variables, the confusion matrix shows matches between ground truth categorical values and output values. For string variables, a string match is displayed. In anonymization tasks, a character-wise overview of truly and falsely redacted characters is given, both as a global metric and for each patient identifier individually. Additionally, each report is listed below the metrics overview. Users can individually select and review each report to double-check and compare the LLM output with the original text and ground truth annotation (**Figure 5B**).

**Figure 4.**
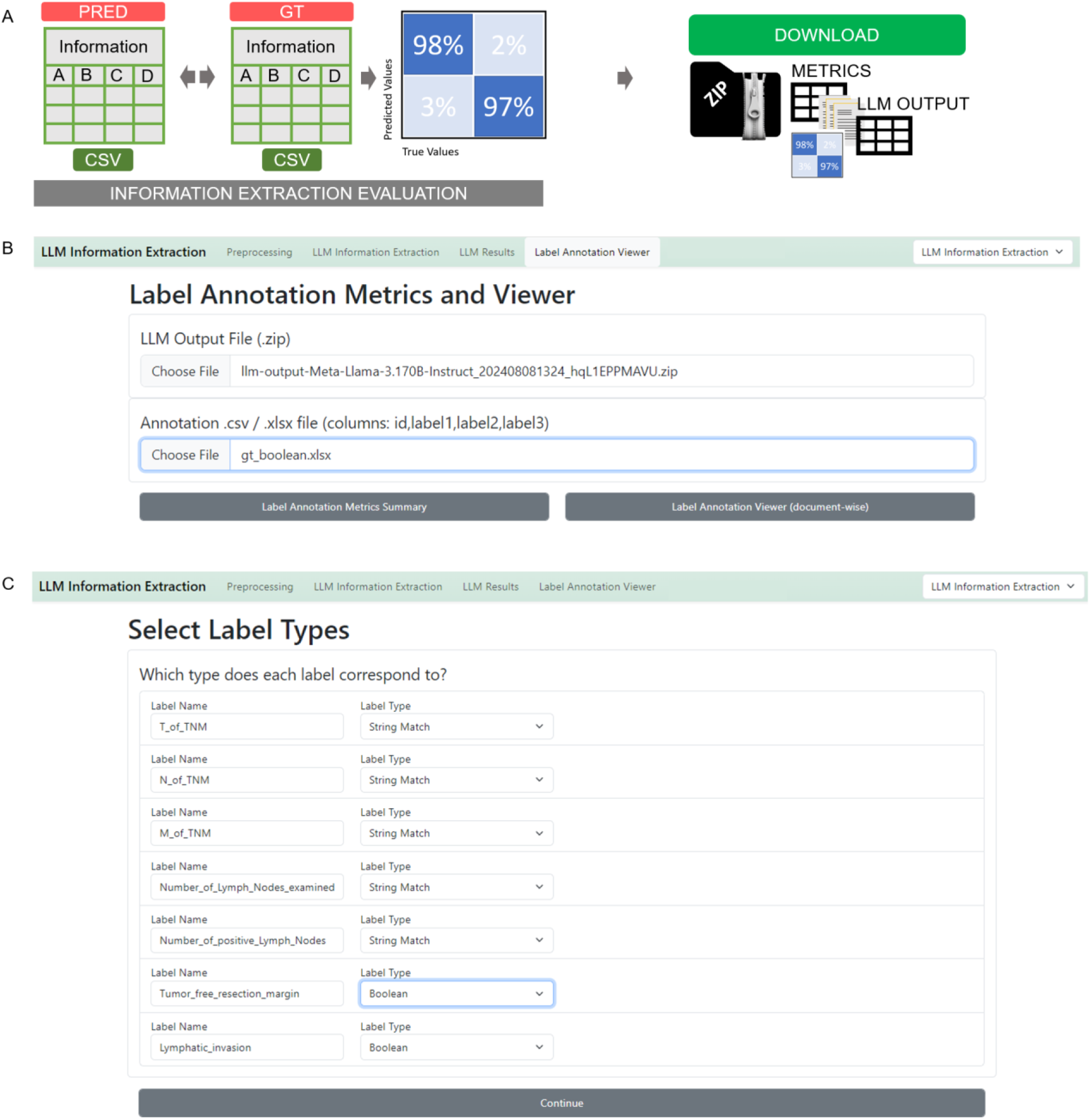
Information extraction evaluation initiation-Stage 4, 11A. **A** Schematic depiction of the evaluation process. LLM output and human made ground truth can be compared automatically and metrics as well as confusion matrices will be calculated and displayed. Those can be downloaded in a metrics-zip file which contains metrics, figures, original documents and LLM output. **B** The LLM Annotation Metrics and Viewer requires upload of the LLM output file (zipfile) and the annotated file (CSV or EXCEL). After initiating the comparison of output file and ground truth file with the “Label Annotation Metrics Summary” button, the label data types of the extracted information variables need to be confirmed. (**C**)

**Figure 5.**
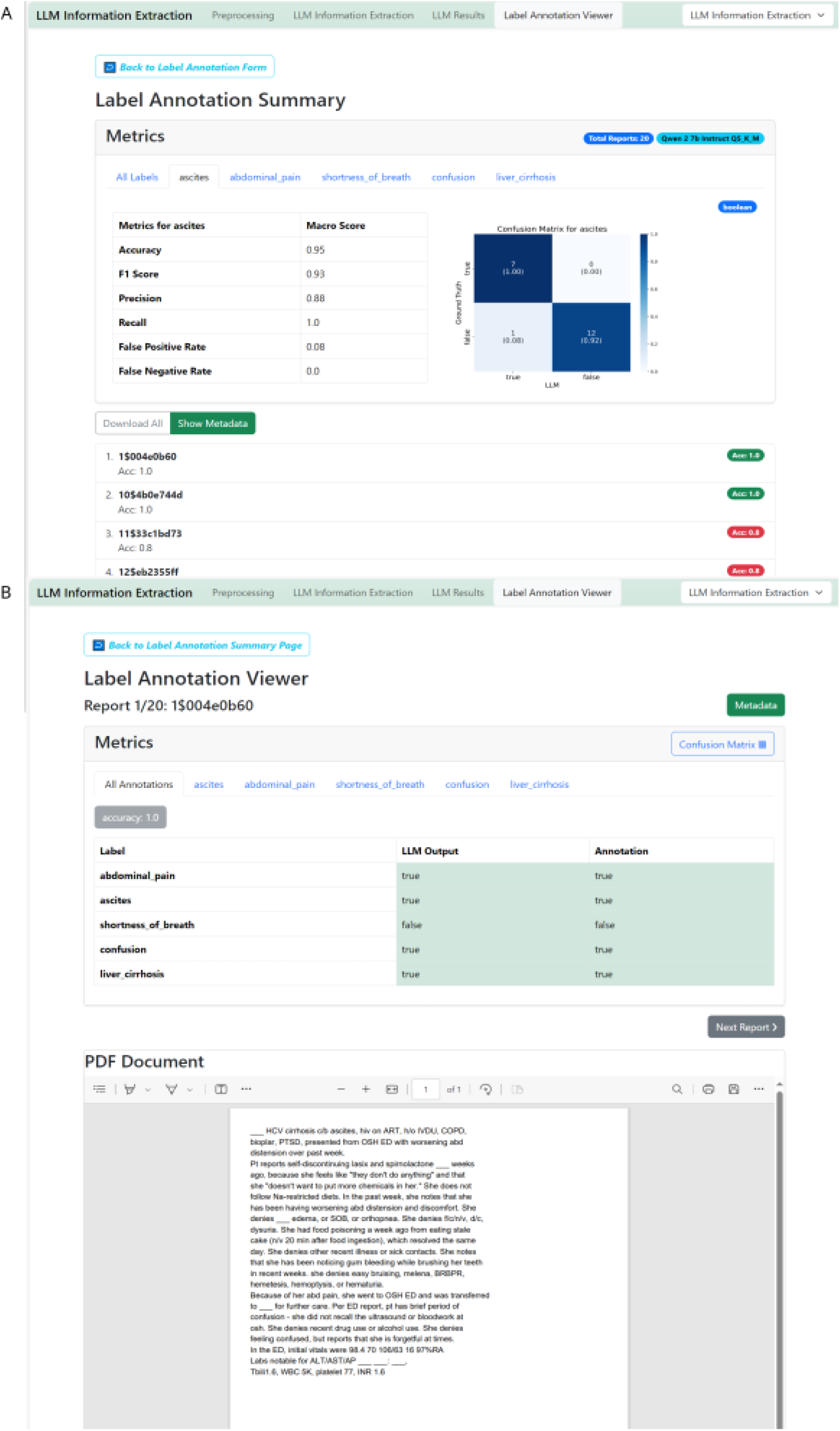
Evaluation of Information Extraction Results GUI, Stage 3, 9-11. **A** Upload the output zip file and the ground truth file. IDs have to match with the document IDs for preprocessing and feature labels have to match the feature names defined for processing. Start the evaluation process by clicking “Label Annotation Metrics Summary”. **B** The label annotation summary provides global metrics accuracy and F1 score, but also allows to check each document individually for all features as well as each feature individually. **C** Clicking on the respective report displays the original text and the model output as well as respective ground truth provided. This enables a seamless evaluation and document revision process.

## Timing

The time required to complete this protocol varies depending on the size of the dataset, the LLM’s size and the available computational resources. The most time-intensive steps are Step 9 “***Prepare the LLM based information extraction”,*** 10 “***Run the LLM based information extraction”*** and Step 12”***Revise Metrics and Files***”. The time estimates provided in **Table 1** are based on running the protocol on the example dataset (comprising n=100 TCGA pathology reports with Llama 3.1 70B model) using a NVIDIA RTX A6000 GPU on a Windows workstation.

**Table 1.** Short description of Information Extraction procedure.

| Step | Name | Timing for TCGA report example |
| --- | --- | --- |
| **Stage 1: Problem definition and data preparations** |  |  |
| 1 | Define the use case | predefined |
| 2 | Assess the input data | predefined |
| 3 | Define the validation strategy | predefined |
| 4 | Prepare the ground truth data | predefined |
| 5 | Download the desired LLMs | ~20 min |
| 6 | Setup for running the pipeline | ~2 min |
| **Stage 2: Data preprocessing** |  |  |
| 7 | Pipeline initiation | ~1 min |
| 8 | Preprocess the data | ~5 min |
| **Stage 3: LLM-based information extraction** |  |  |
| 9 | Prepare the LLM based information extraction | ~5 min |
| 10 | Run the LLM based Information Extraction | ~60 min |
| **Stage 4: Output evaluation** |  |  |
| 11 | A Run evaluation in Information Extraction Mode<br>B Run Evaluation in Anonymizer Mode | ~5 min |
| 12 | Revise Metrics and Files | ~60 min |

## Anticipated Results

Our protocol has demonstrated its efficacy and versatility through application to diverse datasets, notably including the MIMIC dataset^17^ and for psychiatric^16^ and endoscopy report analysis. We demonstrate the pipeline results for both anonymization and IE of eight fictitious clinical letters for patients with pulmonary embolism. Llama-3 70B correctly identified all patients’ first name and last name, gender, age, date of birth and patient id which led to correct redaction of this information in the reports. The character-wise evaluation results in 99.9% specificity and 100% sensitivity for the anonymization of 8 clinical letters, with 98.2% precision for full name redaction, 94.3% precision for first name and 90% for last name redaction (**Table 2**). The discrepancy of IE for all personal identifiers (100% precision and sensitivity) and the redaction metrics is because one fictitious patient’s last name matched the provider’s last name (“Miller”, see **Supplement**) and was therefore also redacted in the exact string match redaction from our pipeline.

**Table 2.** Results LLM-Anonymizer (Macro-Averages)

|  | All labels | Patient name | First name | Last name | Sex | Patient id | Age | Date of birth |
| --- | --- | --- | --- | --- | --- | --- | --- | --- |
| Accuracy in % | 99.9 | 99.9 | 99.9 | 99.8 | 100 | 100 | 99.9 | 100 |
| F1 | 0.994 | 0.988 | 0.969 | 0.944 | 1.0 | 1.0 | 0.958 | 1.0 |
| Precision in % | 98.9 | 97.7 | 94.3 | 90.0 | 100 | 100 | 93.8 | 100 |
| Recall in % | 100 | 100 | 100 | 100 | 100 | 100 | 100 | 100 |
| FPR | 0.0008 | 0.001 | 0.0013 | 0.0026 | 0.0 | 0.0 | 0.0001 | 0.0 |
| FNR | 0.0 | 0.0 | 0.0 | 0.0 | 0.0 | 0.0 | 0.0 | 0.0 |

The fictitious clinical letters report about the clinical course of patients with pulmonary embolism, all of them having different constellations of etiology and risk profiles for this disease. Therefore, we aimed at extracting the presence of leading symptoms such as shortness of breath, chest pain, leg pain or swelling, heart palpitations, cough and dizziness from the clinical letters (**Table 3**). Additionally, information about the embolism side (left, right or bilateral) should be extracted. All symptoms’ presence was correctly identified with 100% precision and sensitivity, except for the symptom “heart palpitations”, which was missed in one clinical letter.

**Table 3.** Results Information Extraction Pulmonary Embolism on Fictitious Reports.

|  | Shortness of breath | Chest pain | Leg pain or swelling | Heart palpitations | Cough | Dizziness | Embolism side |
| --- | --- | --- | --- | --- | --- | --- | --- |
| Accuracy in % | 100 | 100 | 100 | 88.0 | 100 | 100 | 100 |
| F1 | 1.0 | 1.0 | 1.0 | 0.8 | 1.0 | 1.0 | 1.0 |
| Precision in % | 100 | 100 | 100 | 100 | 100 | 100 | 100 |
| Recall in % | 100 | 100 | 100 | 67.0 | 100 | 100 | 100 |
| FPR | 0.0 | 0.0 | 0.0 | 0.0 | 0.0 | 0.0 | 0.0 |
| FNR | 0.0 | 0.0 | 0.0 | 0.33 | 0.0 | 0.0 | 0.0 |

For demonstrating the capacity of our pipeline, we additionally used a dataset previously used for IE with n=100 TCGA pathology reports of patients with colorectal cancer, aiming at extracting information about TNM-stage.^60^ We ran the experiment with Llama 3.1 70B parameter model and achieved an overall accuracy across all variables extracted of 87%. Extracting the T-stage was accurate in 89% of the reports (F1 Score 0.57, Precision 52%, Recall 68%). The N-stage was accurately extracted in 92% of all reports (F1 Score 0.86, Precision 85%, Recall 87%). The M-stage was accurately extracted in 82% (F1 Score 0.69, Precision 68%, Recall 93%). The number of lymph nodes examined was correctly extracted in 87%. The number of lymph nodes positive for cancer cells was correctly extracted in 90%. Whether the resection margin was tumor free could be identified with an accuracy of 86% (F1 Score 0.92, Precision 87%, Recall 99%, FPR93%, FNR 1%). The extraction of whether lymphatic invasion was present or not achieved an accuracy of 86% (F1 Score 0.82, Precision 70%, Recall 100%, FPR 21%, FNR 0%).

A report-by-report error analysis revealed an erroneous ground truth for some reports . When correcting the mistaken ground truth, overall accuracy increased to 88%, 90% for T-stage, 92% for N-stage, 82% for M-stage, 87% for the lymph nodes examined, 90% for cancer-positive lymph nodes, 91% for the tumor free resection margin and 87% for lymphatic invasion (All metrics are shown in **Table 2**). In some cases, the original report contained conflicting information, e.g. it was described: “resection margin negative (carcinoma less than 1mm from the radial margin)” which is in fact oftentimes defined as “resection margin positive”, because there is a high risk of tumor recurrence.^61–63^ Another example was that “M0 Mx” was supposedly accidentally stated at the same time and “M0” extracted by the LLM. In four cases, the error occurred from wrong OCR recognition of the low-quality scans or handwriting within the report. The LLM identified the right characters, which were already incorrectly stored in the document by OCR (e.g. “N1” was recognized as “N:”, “MA” was recognized as “M1” and the roman numbers “I/IV” for positive lymph nodes were recognized as “1/9” and cited by the LLM as such). We repeated preprocessing of the documents and forced OCR with a different OCR engine, “Surya” and found a slight increase in performance metrics (**Table 4**). Detailed guides through the experiments with screenshots, also containing prompts and grammar used, are given in the **Supplement.**

**Table 4.** Results after adapting the erroneous ground truth and adjusting the categories.

|  | All | T stage | N stage | M stage | Number of lymphnodes examined | Number of positive lymphnodes | Tumor free resection margin | Lymphatic invasion |
| --- | --- | --- | --- | --- | --- | --- | --- | --- |
| Data type |  | categorical | categorical | categorical | number | number | boolean | boolean |
| Accuracy in % | 88.0 | 90 | 92 | 82 | 87 | 90 | 91 | 87 |
| F1 |  | 0.57 | .86 | 0.69 |  |  | .95 | 0.85 |
| Precision in % |  | 52 | 85 | 68 |  |  | 92 | 77 |
| Recall in % |  | 68 | 87 | 93 |  |  | 99 | 95 |
| FPR |  |  |  |  |  |  | 0.89 | 0.18 |
| FNR |  |  |  |  |  |  | 0.01 | 0.05 |

**Table 5.** Results after adapted OCR method (output-categories and boolean) Standard Prompt: “You are a helpful medical assistant. You are supposed to extract information from a pathology report from a patient with colorectal cancer. I need to know the TNM stage of the patient. This is a system to describe the amount and spread of cancer in a patient’s body, using TNM. T describes the size of the tumor and any spread of cancer into nearby tissue; N describes spread of cancer to nearby lymph nodes; and M describes metastasis (spread of cancer to other parts of the body). If you find no information about the T, N or M stage, give Tx, Nx or Mx, respectively. If there is “pT1” or “pN”, just skip the “p” and give “T1” etc. Additionally, I need information about the number of lymph nodes examined and the number of positive lymph nodes. Let me know if the resection margin was tumor free and if there was lymphatic invasion. If you do not find information about resection margin or lymphatic invasion, say “not mentioned”. This is the report: {report} ”

|  | All | T stage | N stage | M stage | Number of lymphnodes examined | Number of positive lymphnodes | Tumor free resection margin | Lymphatic invasion |
| --- | --- | --- | --- | --- | --- | --- | --- | --- |
| Accuracy in % | 89 | 90 | 92 | 88 | 88 | 91 | 91 | 88 |
| F1 |  | 0.6 | 0.86 | 0.74 |  |  | 0.95 | 0.86 |
| Precision in % |  | 58 | 85 | 70 |  |  | 93 | 81 |
| Recall in % |  | 67 | 87 | 96 |  |  | 98 | 93 |
| FPR |  |  |  |  |  |  | 0.88 | 0.15 |
| FNR |  |  |  |  |  |  | 0.02 | 0.07 |

These examples demonstrate the successful application of the LLM-AIx-Pipeline for clinical research questions and use cases. It facilitates extracting information from unstructured medical reports with LLMs, thus enabling a structured downstream processing of relevant healthcare data.

## Troubleshooting

| Step | Problem | Possible Reason | Solution |
| --- | --- | --- | --- |
| Data preprocessing |  |  |  |
| 6 | Expecting xx lines in, saw yy | Erroneous CSV, wrong encoding | Make sure you store the data in proper CSV files and ensure UTF-8 coding. |
| LLM-based information extraction |  |  |  |
| 10 | Model could not be loaded on Server | GPU capacity exceeded, too many processes are running in parallel or model is too big for hardware resources | Terminate other processes running on the GPU. If model size is the problem, use a smaller model or higher quantization. |
| 10 | LLM processing very slow | Pipeline is run on CPU of consumer hardware | Loading the LLM to a gPU, if available, increases the speed of the process. Default model upload in the pipeline is GPU upload. |
| 10 | Empty LLM output | LLM output is invalid JSON because the model did not answer anything, did respond with invalid json or the answer was cut off because n_predict is too low. | Increase n_predict. Test with less complex grammar, prompt and other models. |
| 10 | Empty LLM output | N-predict might be too low | Increase n_predict. Test with less complex grammar, prompt and other models. |
| Output Evaluation |  |  |  |
| 11 | Mismatch LLM output with annotation | LLM hallucinated<br>**Supplementary Figure 12** | Improve explanation for this variable in prompt |
| 11 | Mismatch LLM output with annotation | Ground truth is wrong | Refine the ground truth through expert discussion |
| 11 | Mismatch LLM output with annotation | Input text data is conflicting. **Supplementary Figure 13, 14** | Refine the ground truth through expert discussion |
| 11 | Mismatch LLM output with annotation | Annotation is coarser than LLM **Supplementary Figure 15** | Refine the ground truth through expert discussion |
| 11 | Mismatch LLM output with annotation | Annotation is more detailed than LLM **Supplementary Figure 16** | Refine the ground truth through expert discussion |
| 11 | Mismatch LLM output with annotation | Classes are not sufficiently distinguishable, e.g. model is unable to distinguish between “no” and “none”. **Supplementary Figure 17** | Classes need to be defined clearly mutually exclusive and commonly exhaustive and close to interpretation of natural language. E.g. If the presence of “lymphatic invasion” is questioned, the answer options are either “yes”, “no” or “not mentioned”. These classes are closer to human language and therefore clearer for an LLM than “yes, no, none”. |
| 11 | Mismatch LLM output with annotation | Wrong OCR due to bad quality PDFs. **Supplementary Figure 18** | If documents contain a high share of hand-written information and PDFs are of very bad quality, text cannot be extracted correctly, the extraction failure therefore arises from data preprocessing and can be overcome by selecting another OCR method for preprocessing. “Surya” surpasses “tesseract” in general and the visionLLM Phi or trOCR are better in detection hand written text. |
| 11 | Mismatch LLM output with annotation | The LLM output is accurate but has minor character discrepancies compared to the annotation. **Supplementary Figure 19** | Desired output needs to be specified as detailed as possible within the prompt and grammar. |
| 11 | Mismatch LLM output with annotation | Information is present but could not be detected by the LLM.<br>**Supplementary Figure 20** | Add a more detailed explanation within the prompt and give few-shot examples. |
| 11 | Mismatch LLM output with annotation | LLM extracts information better than human rater.<br>**Supplementary Figure 21** | Refine the ground truth through expert discussion |
| 11 | Mismatch LLM output with annotation | LLM lacks implicit knowledge. **Supplementary Figure 22** | Desired output and definition needs to be specified as detailed as possible within the prompt. Add few-shot examples. |
| 11 | Mismatch LLM output with annotation | Output categories are misdefined.<br>**Supplementary Figure 23** | Make sure the output categories are the same as in annotated ground truth |

## Supporting information

Manuscript Supplement

## Data availability

All research procedures were conducted in accordance with the Declaration of Helsinki. Ethics approval was granted by the ethics committee of Technical University Dresden, reference number BO-EK-400092023.

## Code availability

The open-source software for the implementation of the IE experiments is available on GitHub under a CC-BY-NC license (https://github.com/KatherLab/LLMAIx).

## Acknowledgements

Thank you to the testers of the protocol, Chiara M.L. Loeffler, Hannah S.Muti, Ariana Bonetti who tested the protocol on several information extraction tasks. Special thanks are extended to Chiara M. L. Loeffler for providing the annotated TCGA dataset and for support in pipeline naming. Appreciation is also expressed to Katherine Hewitt for pathological advice in adjusting the ground truth for TCGA reports.

## Author contributions

ICW conceptualized the methodology and protocol and designed the pipeline with MEL, JNK and FW. FW, MvT and ICW developed the software and wrote the technical documentation with contributions of JZ and MEL. ICW, MEL, FW, DF and HB tested and adapted the software. ICW and MEL interpreted the analyzed data. ICW, MEL, FW, JNK and DF were writing and reviewing the initial manuscript. All authors wrote and reviewed the protocol and approved the final version for submission. MPE and JNK provided supervision and resources for the project.

As per the ICMJE guidelines of April 2023, we hereby disclose that the following tools were used to write this article: Microsoft Word and Google Documents as Word processing software, ChatGPT-4 for checking and correcting spelling and grammar, Midjourney V5.2 for icon generation.

