## Supplementary material for "LLM-AIx: An open source pipeline for Information Extraction from unstructured medical text based on privacy preserving Large Language Models": Manuscript Supplement

#### Guide through anonymization example

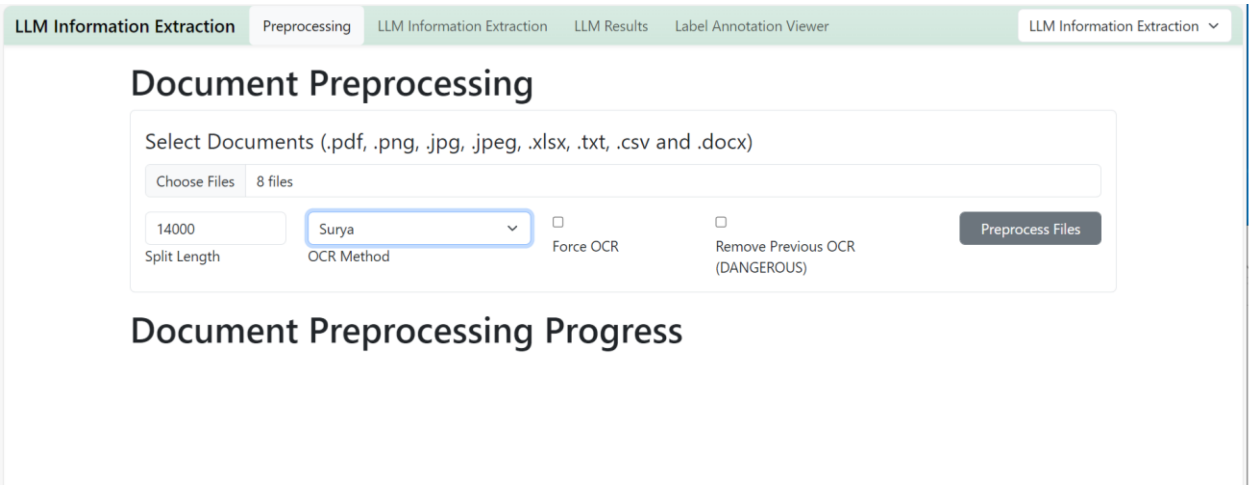


**Supplementary Figure 1 - Preprocessing of fictitious clinical letters**.

Preprocessing is performed equally in the LLM Information Extraction mode and in the Anonymizer mode. The 8 fictitious letters were uploaded and the Optical Character Recognition (OCR) method defined. Since all PDFs already contained text, no OCR was necessary. The preprocessing process can be started by clicking the “Preprocess Files” button.  A progress bar indicates the process and a “Download” button allows downloading the preprocessed files as a zip file.

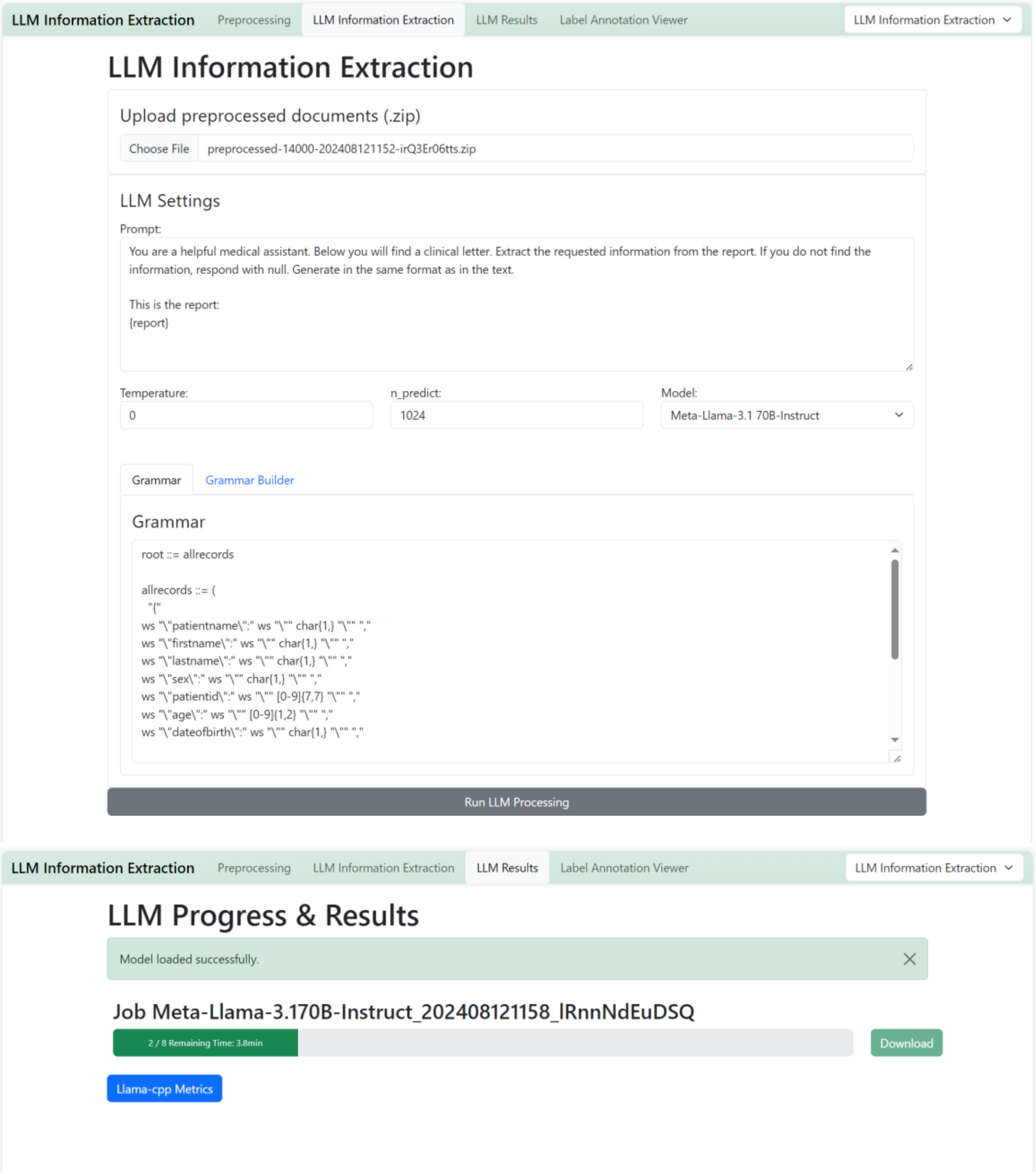


**Supplementary Figure 2 - Preparation of LLM based information extraction.**

**A** The preprocessed zip file was uploaded in the respective field ("Upload preprocessed documents"). Within the LLM settings, the prompt can be adapted. The prompt used for the example anonymization task is given here. The hyperparameter temperature was set to zero to ensure the most deterministic LLM output and the model chosen was Meta's Llama 3.1 70B in 4-bit quantization and GGUF format available from "huggingface". With the grammar builder, the shown grammar could be defined to ensure consistent JSON formatted output with the desired variables. The button "Run LLM Processing" allows to start the LLM based information extraction. **B** The correct model loading is indicated with a green bar stating "Model loaded successfully". A progress bar displays the remaining process time and a "Download" button allows downloading the output zip file as soon as the process is finished. This process can be performed in both Information extraction and Anonymizer mode.

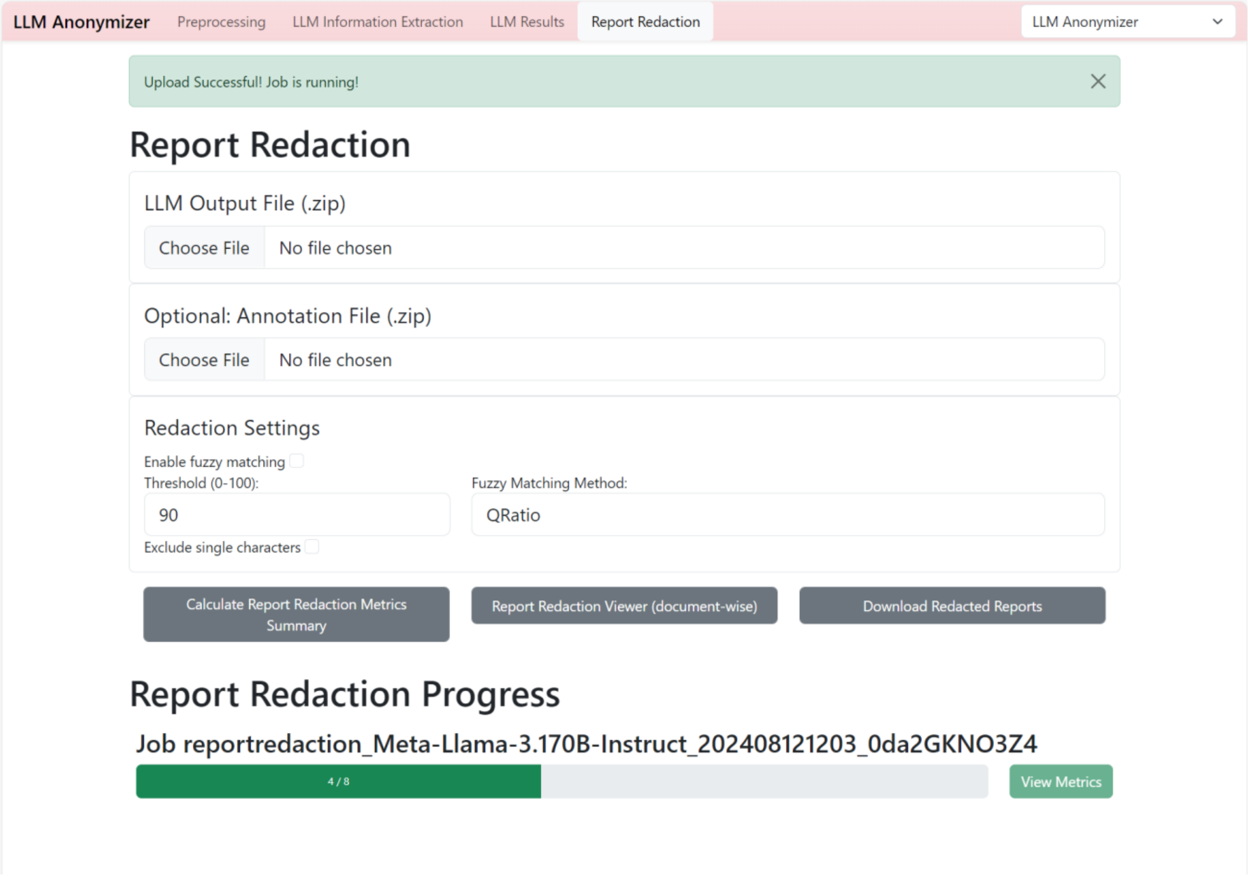


**Supplementary Figure 3 - Report Redaction.**

The analysis and report redaction part is specific to the Anonymizer mode, therefore the mode needs to be switched in the right upper corner of the window. The LLM output zip file was downloaded for the 8 fictitious clinical letters as well as the annotation file (human annotations were performed with the annotation tool “Inception” and downloaded as JSON files. These JSON files were zipped and uploaded). The extracted identifiers were now redacted within the original clinical letters with exact character matching. The pipeline additionally allows for “fuzzy matching”, a technique used to find strings that are approximately rather than exactly equal. This can be useful for typographical errors or variations in spelling. Two options are available: Qratio and Wratio. Qratio compares two strings and provides a similarity score between 0 and 100, zero meaning that the two strings are completely different. Wratio adjusts the basic Qratio by string length and variations. The threshold can be flexibly set based on the user’s needs between 0 (everything matches, regardless of similarity) and 100 (only exact string matches are considered).

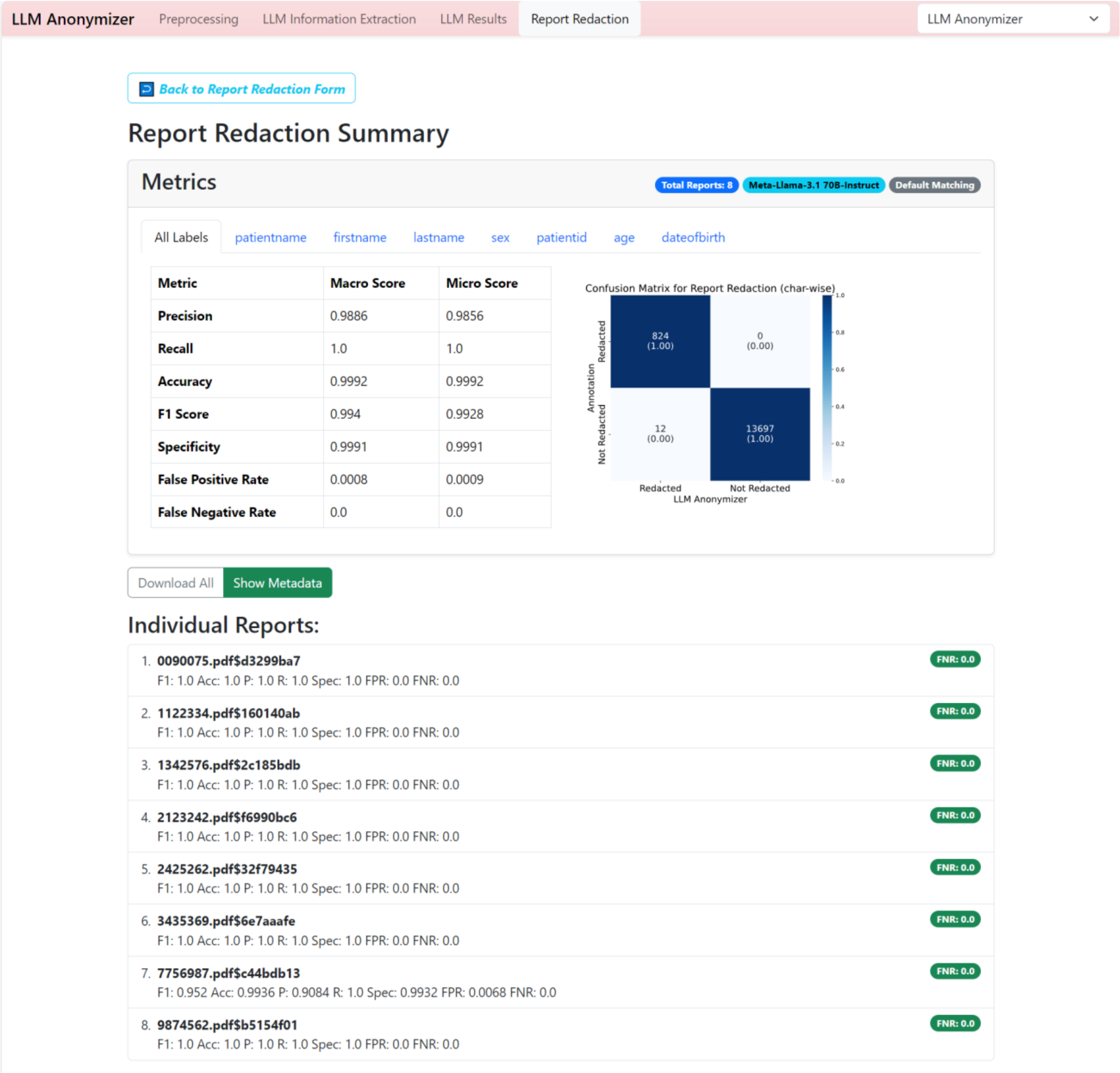


**Supplementary Figure 4 - Report Redaction Summary.**

The report redaction summary displays character-wise matching metrics between the human annotations and the pipeline-redacted documents. Macro and Micro scores are shown for all labels as well as for each variable extracted. Blue and grey tags indicate the total number of reports processed (8 fictitious clinical letters), the model used (Meta-Llama 3.1 70B) and the matching algorithm (Default: Exact match). Additionally, false negative rates (FNR) are displayed for all clinical letters individually. These documents can be opened and reviewed by clicking on each document in the list.

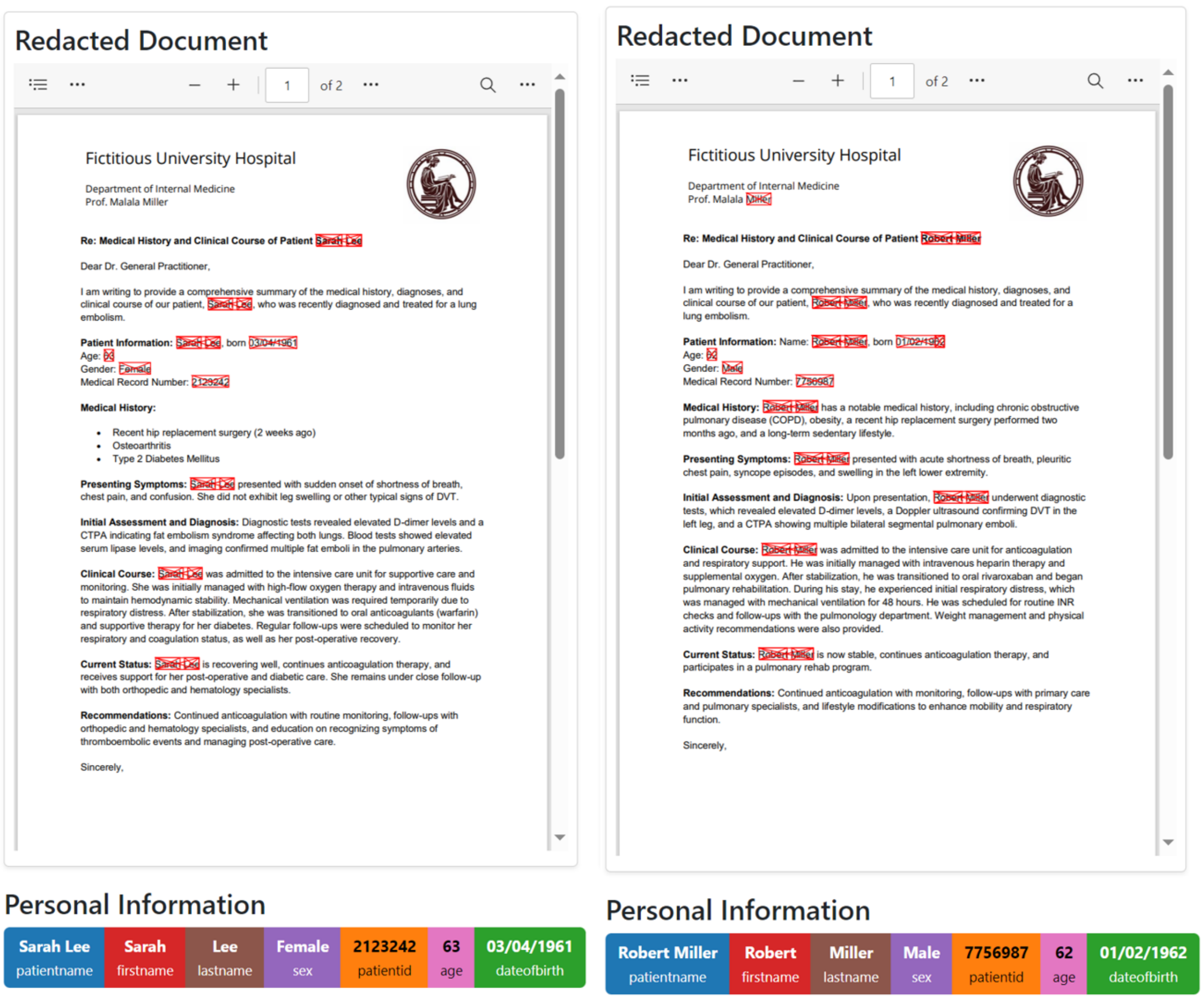


**Supplementary Figure 5 - Redacted document view.**

Each report can be reviewed individually. Two examples from Anonymization example with 8 fictitious clinical letters are shown here. The “Personal Information” bar displays the extracted personal information. The original clinical letter can be reviewed and red boxes indicate The extracted information is then displayed separately and red boxes indicate the fields that are blackened when downloading the redacted documents.

Fictitious University Hospital

Department of Internal Medicine
Prof. Malala Miller

**Re: Medical History and Clinical Course of Patient Elisabeth Zhu**

Dear Dr. General Practitioner,

I am writing to provide a comprehensive summary of the medical history, diagnoses, and clinical course of our patient, Elisabeth Zhu, who was recently diagnosed and treated for a lung embolism.

**Patient Information:** Elisabeth Zhu, dob 07/07/1970
Age: 48
Gender: Female
Medical Record Number: 1342576

**Medical History:** Elisabeth Zhu has a notable medical history, including:

- History of autoimmune disorder (Systemic Lupus Erythematosus)
- Long-term corticosteroid therapy
- Recent minor surgery (cholecystectomy)

**Presenting Symptoms:**
Elisabeth Zhu, a 48-year-old female with a known history of Systemic Lupus Erythematosus (SLE) and long-term corticosteroid therapy, presented with progressive dyspnea over the past week. She described experiencing chest tightness, particularly noticeable during exertion. Additionally, she reported significant fatigue and malaise, impacting her daily activities. On physical examination, there was mild swelling observed in her left leg, raising concerns for a potential thromboembolic event.

**Initial Assessment and Diagnosis:** Diagnostic tests included:

- Elevated D-dimer levels
- Venous Doppler ultrasound confirming DVT in the left leg
- CTPA showing pulmonary embolism in the right upper lobe

**Clinical Course:**

Upon admission to the general medical ward, Elisabeth Zhu was initiated on low molecular weight heparin (LMWH) therapy for anticoagulation and placed under close monitoring for vital signs and oxygen saturation. Subsequently, her management included a transition from LMWH to oral anticoagulants, specifically dabigatran. Concurrently, her corticosteroid therapy was adjusted in consultation with her rheumatologist to manage her Systemic Lupus Erythematosus (SLE) effectively. During her hospitalization, Elisabeth experienced a flare-up of her lupus symptoms, which was managed with appropriate immunosuppressive therapy.

Follow-up care involved regular monitoring of her coagulation status and ongoing management of her lupus. Additionally, she received education on recognizing symptoms of

Fictitious University Hospital

Department of Internal Medicine
Prof. Malala Miller

thromboembolic events to ensure prompt identification and treatment of any future occurrences.

**Current Status:** Elisabeth Zhu is stable, continues anticoagulation therapy, and is under regular follow-up with both rheumatology and hematology specialists.

**Recommendations:**

- Continued anticoagulation with routine monitoring
- Ongoing coordination between rheumatology and hematology for integrated care
- Education on recognizing and preventing thromboembolic events

Sincerely,

F. Finch

Dr. Fictitious Finch

Fictitious University Hospital

Department of Internal Medicine
Prof. Malala Miller

**Re: Medical History and Clinical Course of Patient [redacted]**

Dear Dr. General Practitioner,

I am writing to provide a comprehensive summary of the medical history, diagnoses, and clinical course of our patient, [redacted], who was recently diagnosed and treated for a lung embolism.

**Patient Information:** [redacted], dob [redacted]
Age: [redacted]
Gender: [redacted]
Medical Record Number: [redacted]

**Medical History:** [redacted] has a notable medical history, including:

- History of autoimmune disorder (Systemic Lupus Erythematosus)
- Long-term corticosteroid therapy
- Recent minor surgery (cholecystectomy)

**Presenting Symptoms:**
[redacted], a [redacted]-year-old [redacted] with a known history of Systemic Lupus Erythematosus (SLE) and long-term corticosteroid therapy, presented with progressive dyspnea over the past week. She described experiencing chest tightness, particularly noticeable during exertion. Additionally, she reported significant fatigue and malaise, impacting her daily activities. On physical examination, there was mild swelling observed in her left leg, raising concerns for a potential thromboembolic event.

**Initial Assessment and Diagnosis:** Diagnostic tests included:

- Elevated D-dimer levels
- Venous Doppler ultrasound confirming DVT in the left leg
- CTPA showing pulmonary embolism in the right upper lobe

**Clinical Course:**

Upon admission to the general medical ward, [redacted] was initiated on low molecular weight heparin (LMWH) therapy for anticoagulation and placed under close monitoring for vital signs and oxygen saturation. Subsequently, her management included a transition from LMWH to oral anticoagulants, specifically dabigatran. Concurrently, her corticosteroid therapy was adjusted in consultation with her rheumatologist to manage her Systemic Lupus Erythematosus (SLE) effectively. During her hospitalization, [redacted] experienced a flare-up of her lupus symptoms, which was managed with appropriate immunosuppressive therapy.

Follow-up care involved regular monitoring of her coagulation status and ongoing management of her lupus. Additionally, she received education on recognizing symptoms of

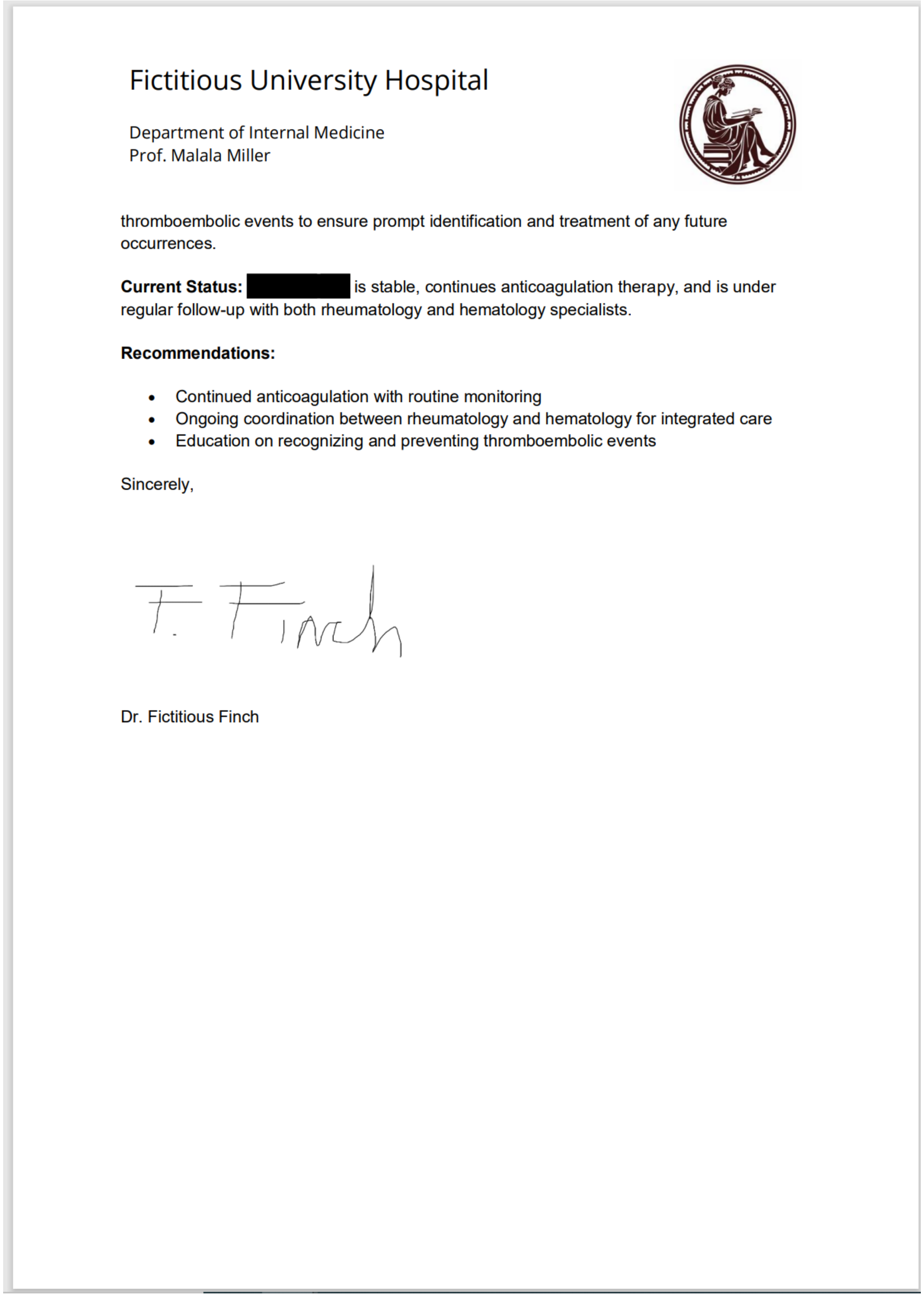

Fictitious University Hospital

Department of Internal Medicine
Prof. Malala Miller

thromboembolic events to ensure prompt identification and treatment of any future occurrences.

**Current Status:** [redacted] is stable, continues anticoagulation therapy, and is under regular follow-up with both rheumatology and hematology specialists.

**Recommendations:**

- Continued anticoagulation with routine monitoring
- Ongoing coordination between rheumatology and hematology for integrated care
- Education on recognizing and preventing thromboembolic events

Sincerely,

F. Finch

Dr. Fictitious Finch

We present the redacted clinical letter from one out of eight fictitious examples here. The downloaded, redacted PDFs contain blackened identifiers as defined by the user. The text layer behind is completely removed.

A

B

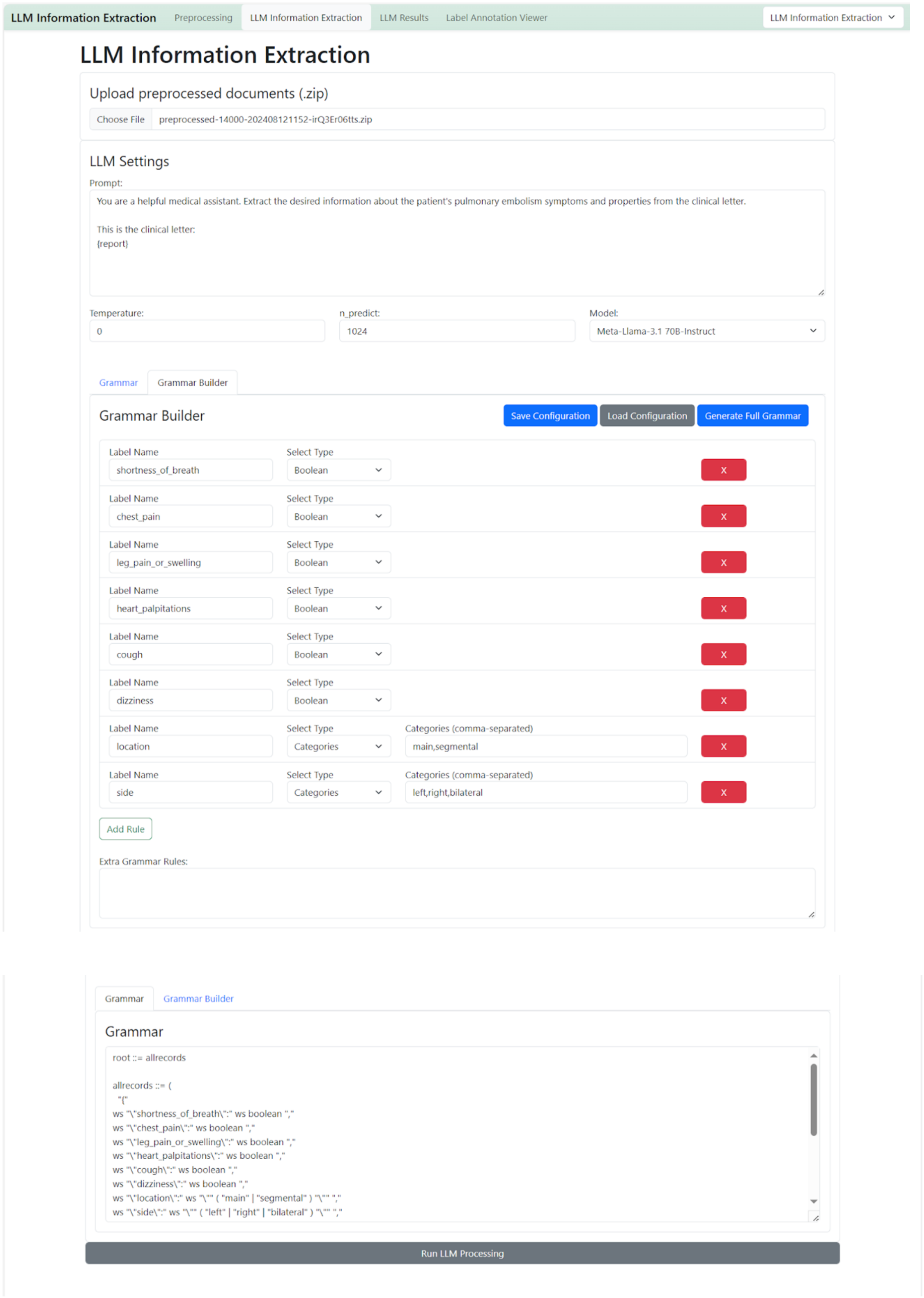

LLM Information Extraction
Preprocessing
LLM Information Extraction
LLM Results
Label Annotation Viewer
LLM Information Extraction
LLM Information Extraction
Upload preprocessed documents (.zip)
Choose File
preprocessed-14000-202408121152-irQ3Er06tts.zip
LLM Settings
Prompt:
You are a helpful medical assistant. Extract the desired information about the patient's pulmonary embolism symptoms and properties from the clinical letter.
This is the clinical letter:
{report}
Temperature:
0
n_predict:
1024
Model:
Meta-Llama-3.1 70B-Instruct
Grammar
Grammar Builder
Grammar Builder
Save Configuration
Load Configuration
Generate Full Grammar
Label Name
shortness_of_breath
Select Type
Boolean
chest_pain
leg_pain_or_swelling
heart_palpitations
cough
dizziness
location
Categories
Categories (comma-separated)
main,segmental
side
left,right,bilateral
X
Add Rule
Extra Grammar Rules:
Grammar
root ::= allrecords
allrecords ::= (
"{"
ws "\"shortness_of_breath\":" ws boolean ","
ws "\"chest_pain\":" ws boolean ","
ws "\"leg_pain_or_swelling\":" ws boolean ","
ws "\"heart_palpitations\":" ws boolean ","
ws "\"cough\":" ws boolean ","
ws "\"dizziness\":" ws boolean ","
ws "\"location\":" ws "\"" ( "main" | "segmental" ) "\"" ","
ws "\"side\":" ws "\"" ( "left" | "right" | "bilateral" ) "\"" ","
Run LLM Processing

**Supplementary Figure 6 - Preparation for LLM based information extraction for pulmonary embolism symptoms.**

We defined a set of symptoms as well as the location of the pulmonary embolism as variables to extract from the 8 fictitious clinical letters and built a grammar with the grammar builder tool (**A**) and then generated and adopted in the “Grammar” field (**B**) . The prompt was very simple and is given in the “Prompt” field. The model used was Meta’s Llama 3.1 70B in 4-bit quantization and GGUF format available from “huggingface”.

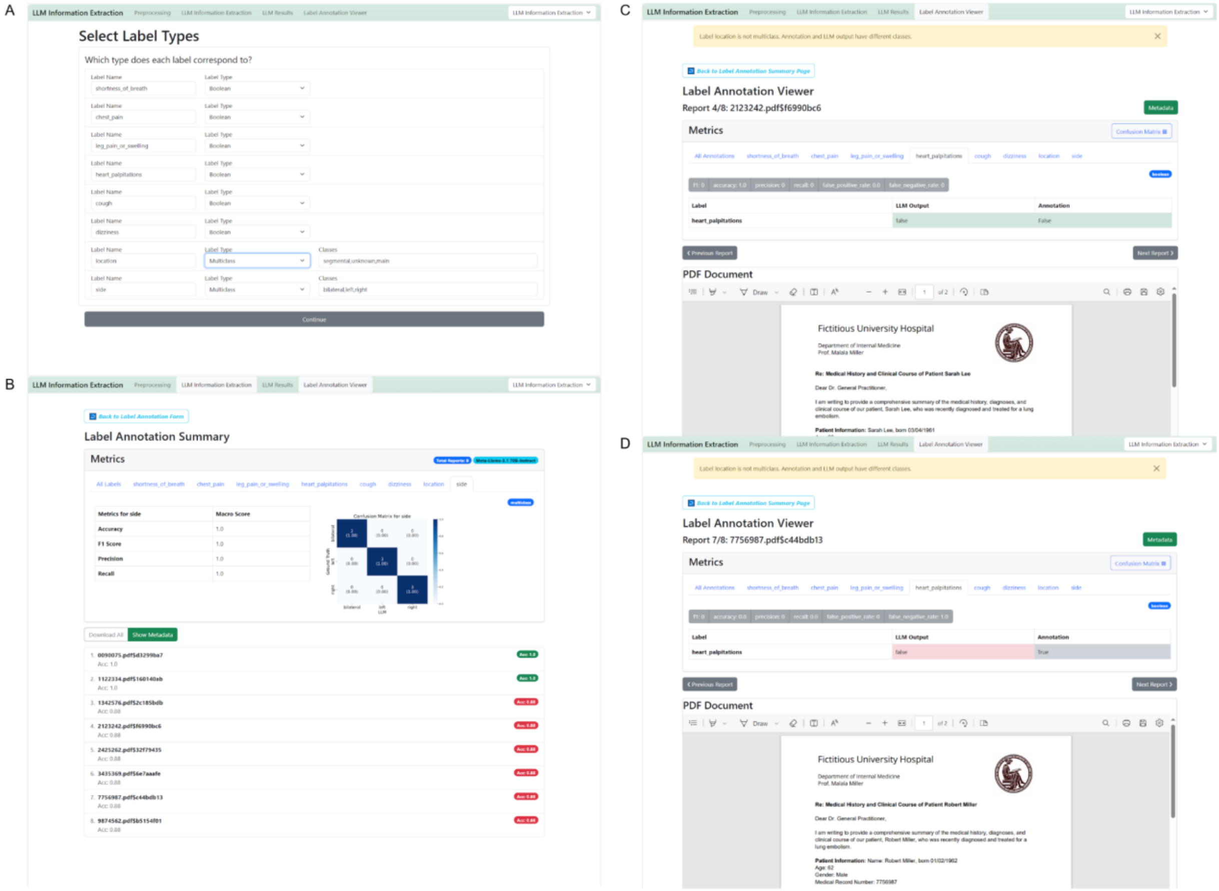


**Supplementary Figure 7 - Evaluation of Information Extraction.**

**A** For information extraction tasks, it is essential to confirm the data types of variables. Boolean variables enable the use of more diverse metrics than string variables, which are limited to simple string matching. When defining multiclass variables, their characteristics should be provided in a comma-separated format.**B** The annotation summary presents metrics for all variables collectively and for each variable individually. For categorical and boolean variable types, confusion matrices are shown alongside a table that includes comprehensive metrics such as accuracy, precision, recall, and F1 score. The results for all variables can be reviewed on a per-document basis. **C** An example of a correctly identified clinical letter, where the patient complains about palpitations. **D** An example of an incorrect output from the language model (LLM) for the symptom of palpitations.

### Guide through experiment 2

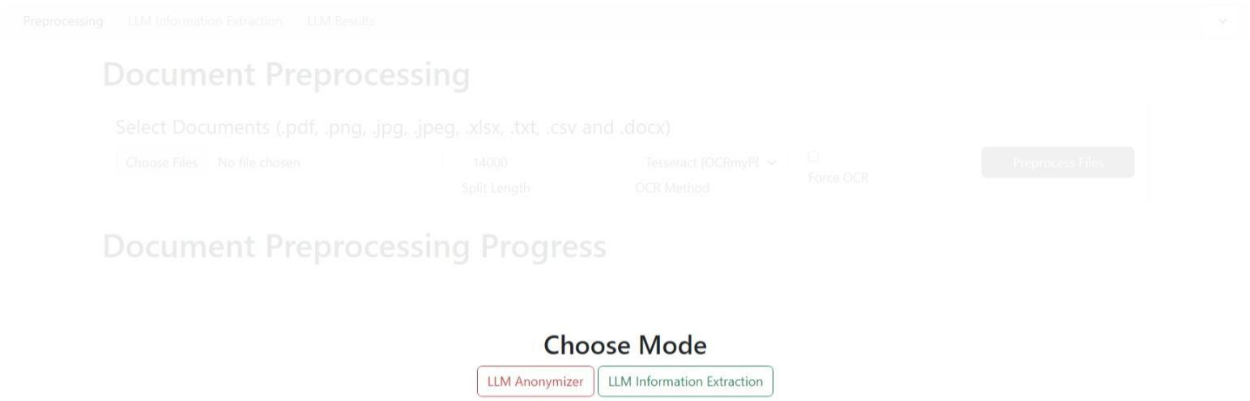


**Supplementary Figure 8** - Choose Mode. Depending on the task, in this case information extraction, the mode can be chosen when starting the process but also switched flexibly during the process.

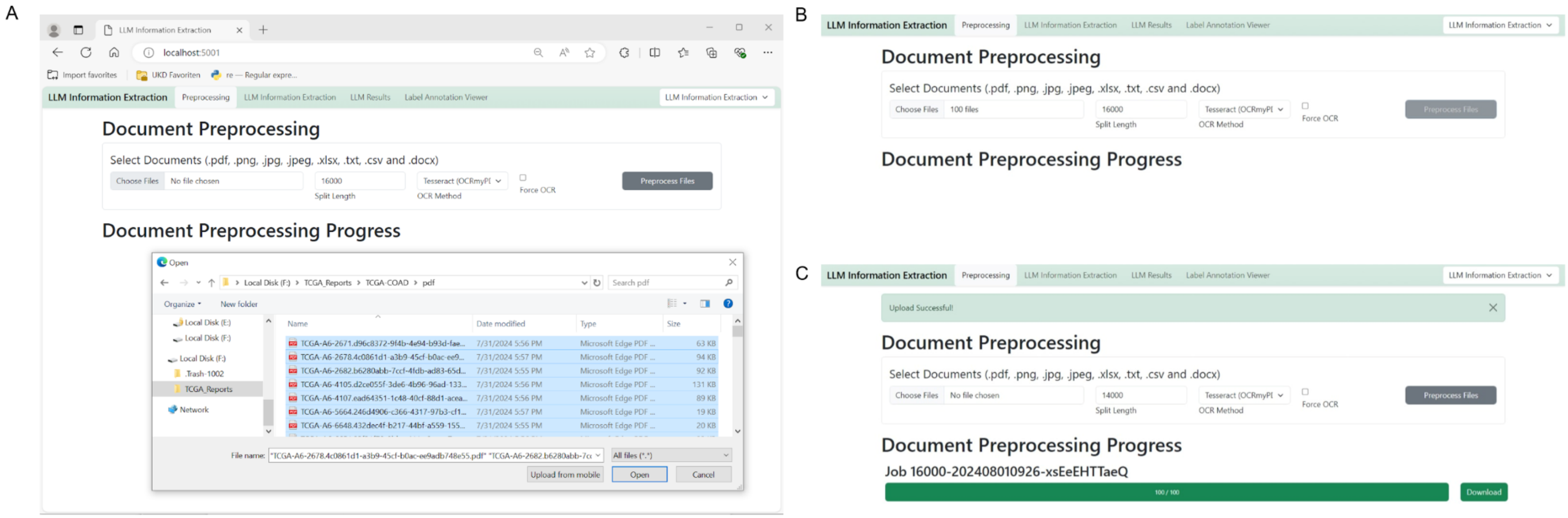


**Supplementary Figure 9 - Data preprocessing for TCGA reports.** All 100 TCGA reports in portable document format (PDF) were uploaded simultaneously. The preprocessing process was initiated with the “Preprocess files” button and a zip file containing the preprocessed documents could be downloaded. The PDFs already contained a text layer, therefore preprocessing without optical character recognition (OCR) took place.

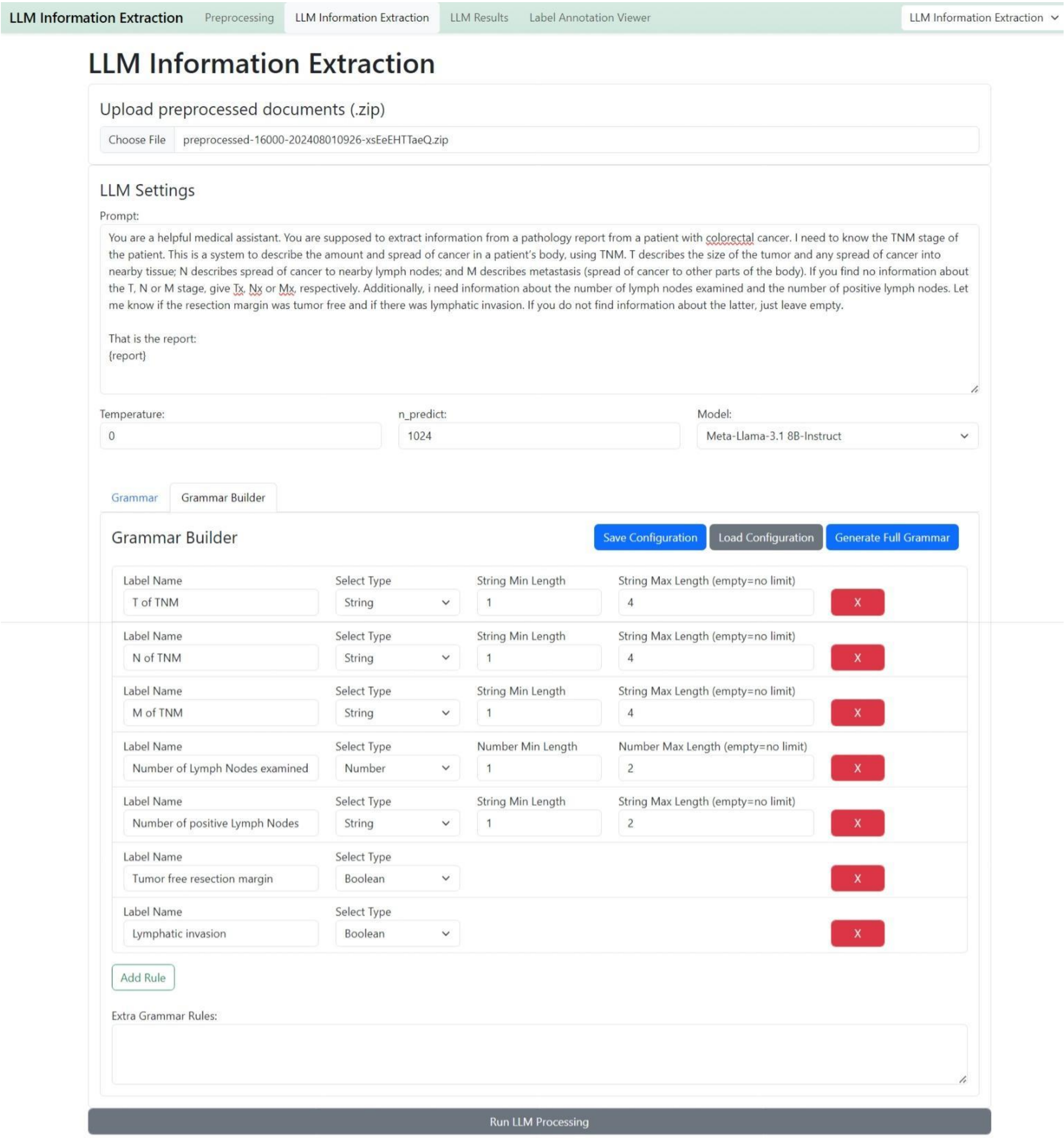


**Supplementary Figure 10 - LLM Information Extraction.**

The preprocessed zip file of all 100 TCGA reports was uploaded. Afterwards, the prompt was defined as specified in the figure and model as well as hyperparameters were chosen. The grammar builder assisted for defining the grammar. Initially, T-, N-, and M-stage were defined as strings, “Number of lymph nodes examined” and “-positive” were defined as numbers and the variables “Tumor free resection margin” and “lymphatic invasion" were defined as boolean variables.

### Troubleshooting Examples

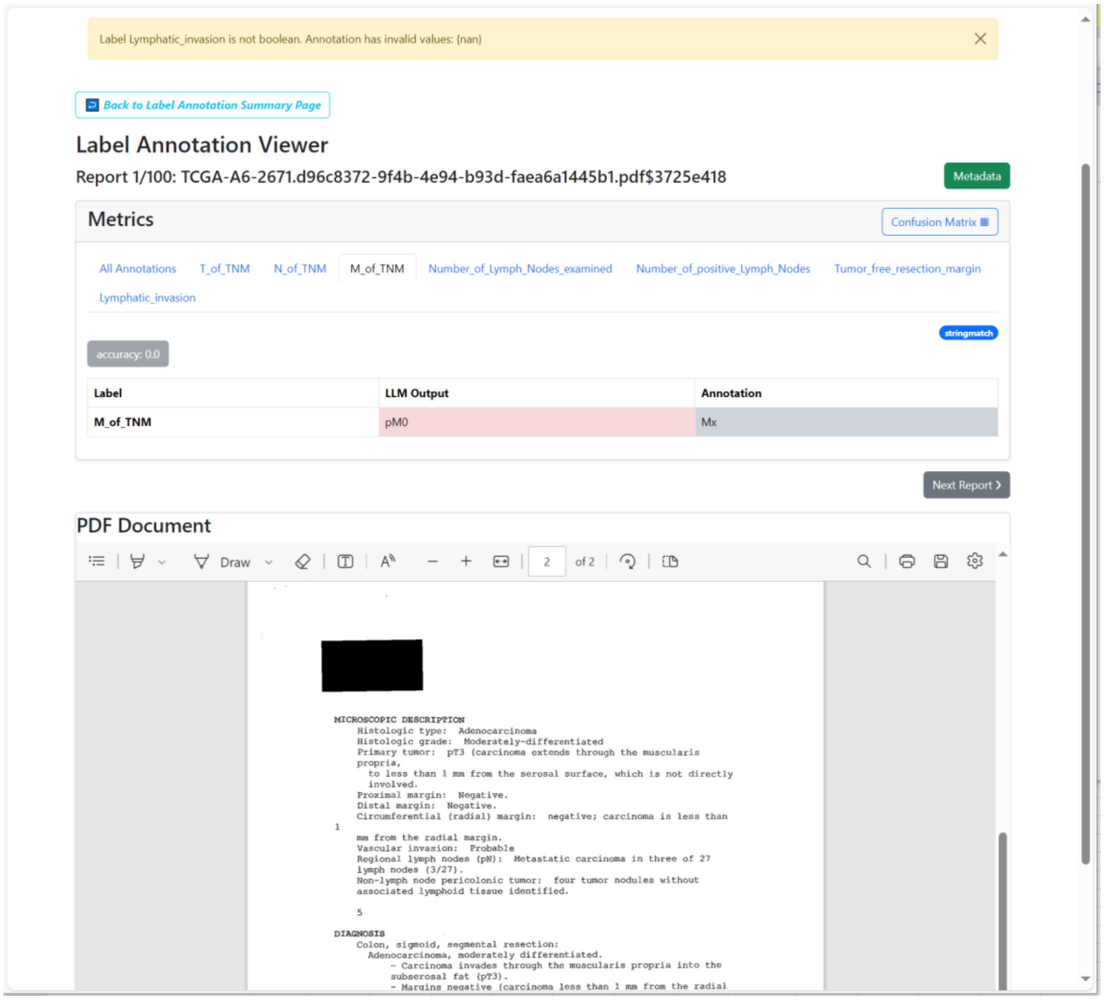


**Supplementary Figure 12 - Model Hallucination.**

There is no information in the pathology report about the metastatic status of the tumor. Therefore, the correct answer would be “Mx”, but the model outputs pM0.

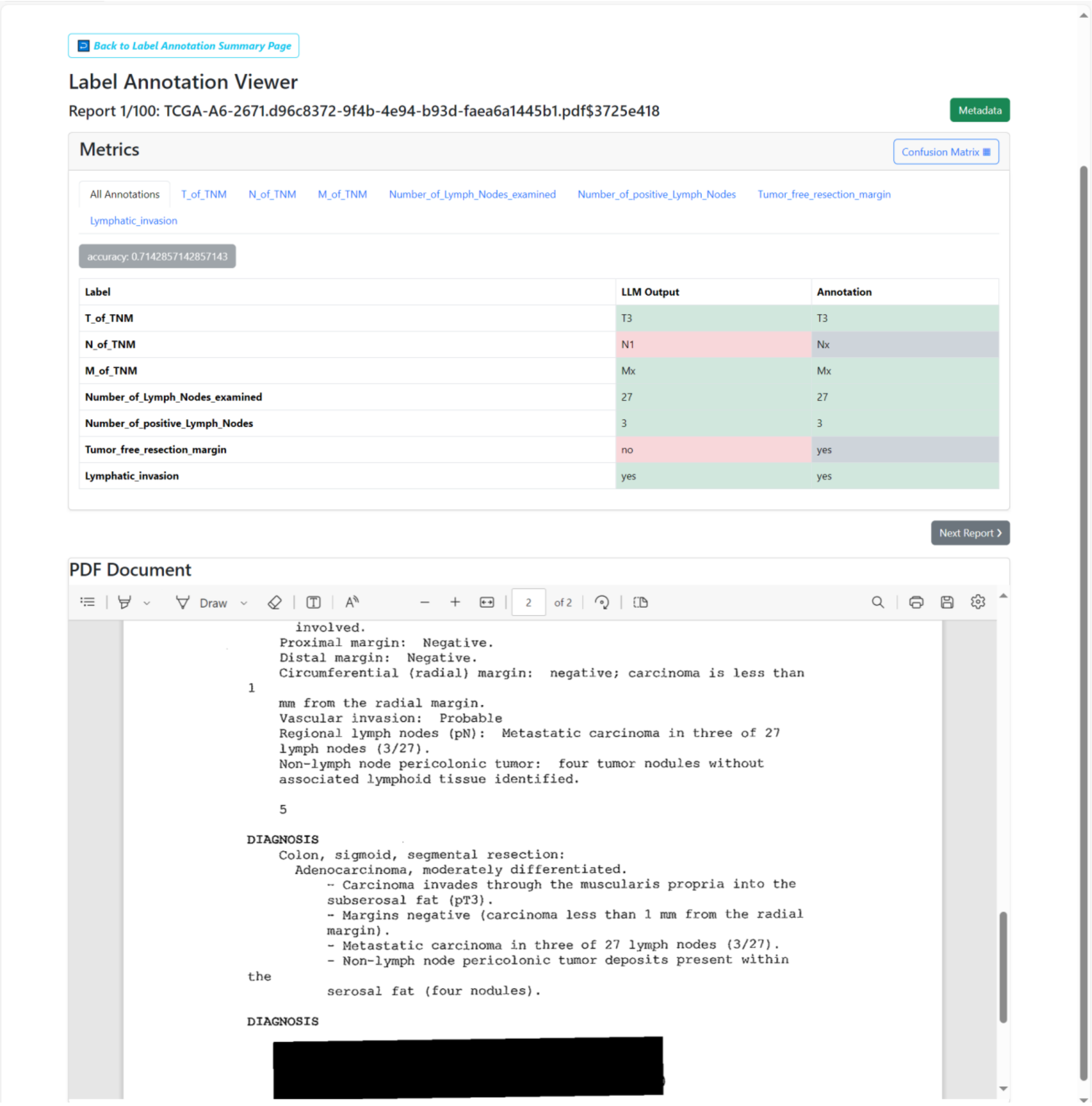


**Supplementary Figure 13 - Conflicting input text data, Example 1.**

For colorectal cancer, negative margins in a pathology report refer to the fact that the resection margins are tumor-free. The report states that "margins [are] negative", but if the carcinoma is located less than 1mm distant from the resection margin, this is accompanied by bad prognosis and therefore could be considered as "margins positive". In this case, a consensus needed to be made in order to define the absolute ground truth.

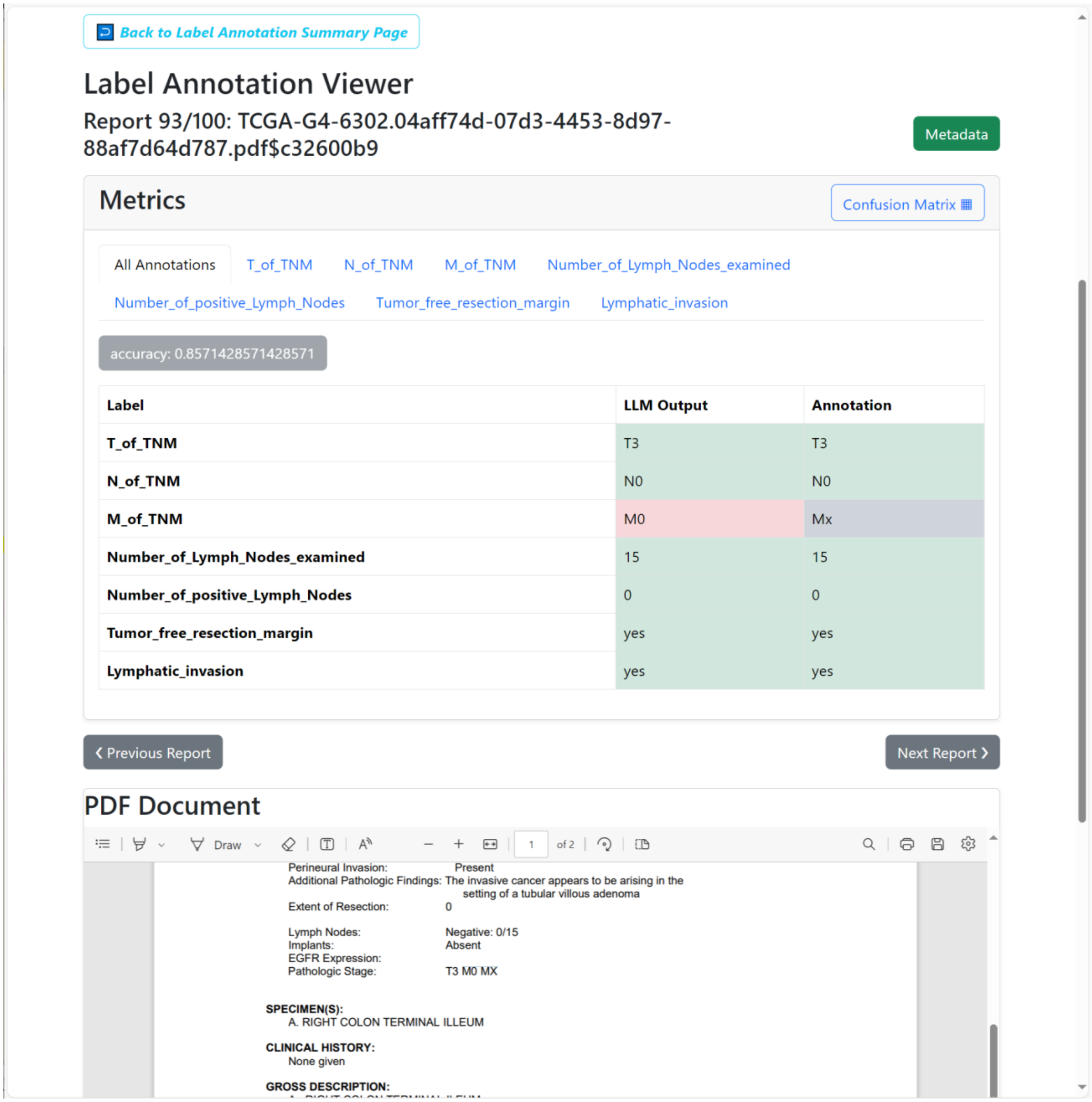

Back to Label Annotation Summary Page

#### Label Annotation Viewer

Report 93/100: TCGA-G4-6302.04aff74d-07d3-4453-8d97-88af7d64d787.pdf$c32600b9

Metadata

##### Metrics

Confusion Matrix

All Annotations T_of_TNM N_of_TNM M_of_TNM Number_of_Lymph_Nodes_examined Number_of_positive_Lymph_Nodes Tumor_free_resection_margin Lymphatic_invasion

accuracy: 0.8571428571428571

| Label | LLM Output | Annotation |
|---|---|---|
| T_of_TNM | T3 | T3 |
| N_of_TNM | N0 | N0 |
| M_of_TNM | M0 | Mx |
| Number_of_Lymph_Nodes_examined | 15 | 15 |
| Number_of_positive_Lymph_Nodes | 0 | 0 |
| Tumor_free_resection_margin | yes | yes |
| Lymphatic_invasion | yes | yes |

Previous Report Next Report

##### PDF Document

Draw 1 of 2

Perineural Invasion: Present
Additional Pathologic Findings: The invasive cancer appears to be arising in the setting of a tubular villous adenoma
Extent of Resection: 0

Lymph Nodes: Negative: 0/15
Implants: Absent
EGFR Expression:
Pathologic Stage: T3 M0 MX

SPECIMEN(S):
A. RIGHT COLON TERMINAL ILLEUM

CLINICAL HISTORY:
None given

GROSS DESCRIPTION:

**Supplementary Figure 14 - Conflicting Input text data, Example 2**.

The pathologic stage section gives conflicting information about the metastasis status by stating “T3 M0 Mx”. In this report, the only specimen examined is colorectal tumor tissue, there is no information about metastasis, therefore MX should be the correct answer. The LLM extracts “M0”, which is also conflictingly stated in the document.

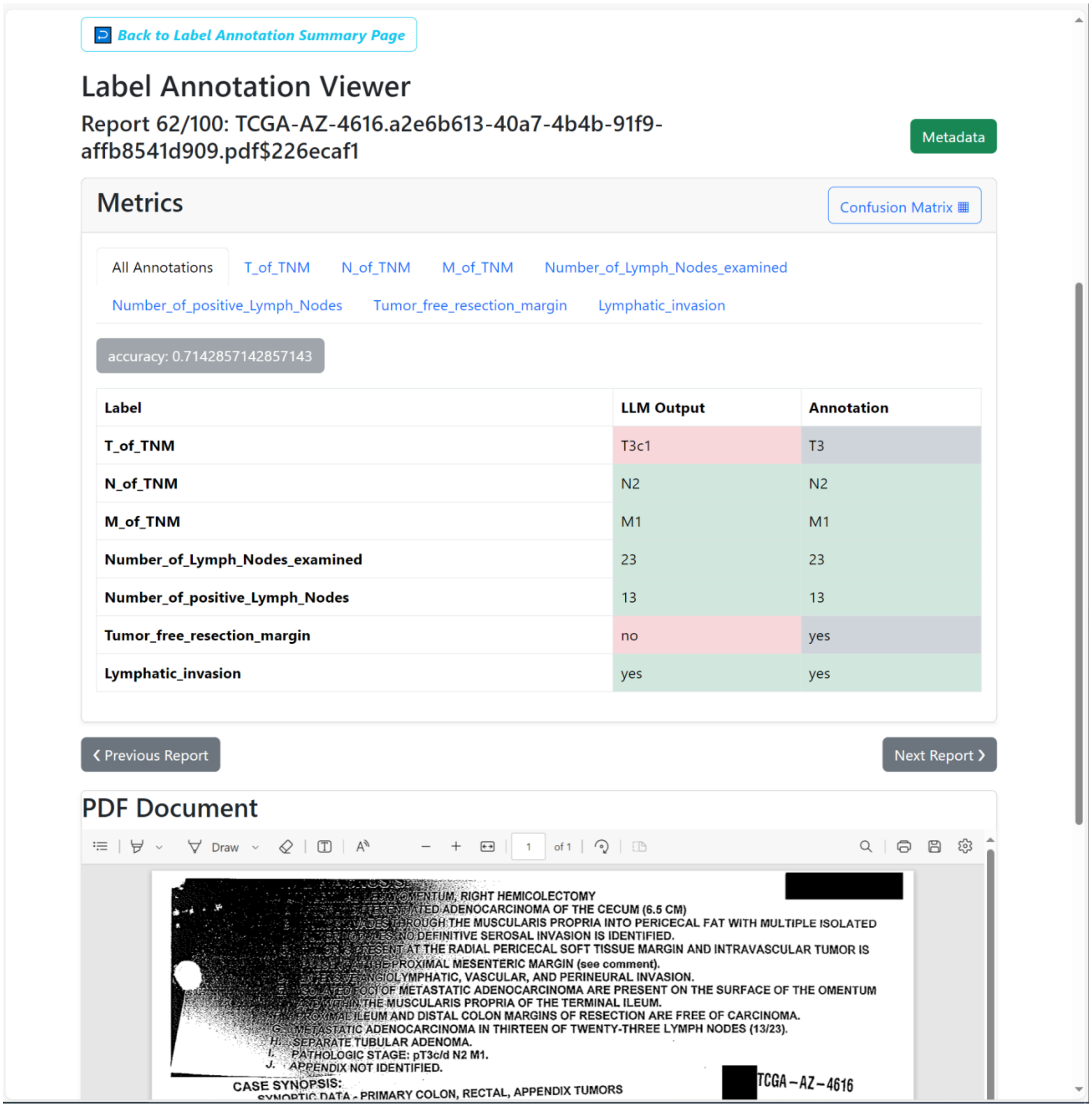


**Supplementary Figure 15 - Annotation is coarser than LLM.**

The LLm extracts the described T-stage from the pathology report, which is “T3c1”, which is more detailed than the annotation which only provides “T3”, therefore detected as mismatching, even though both are correct.

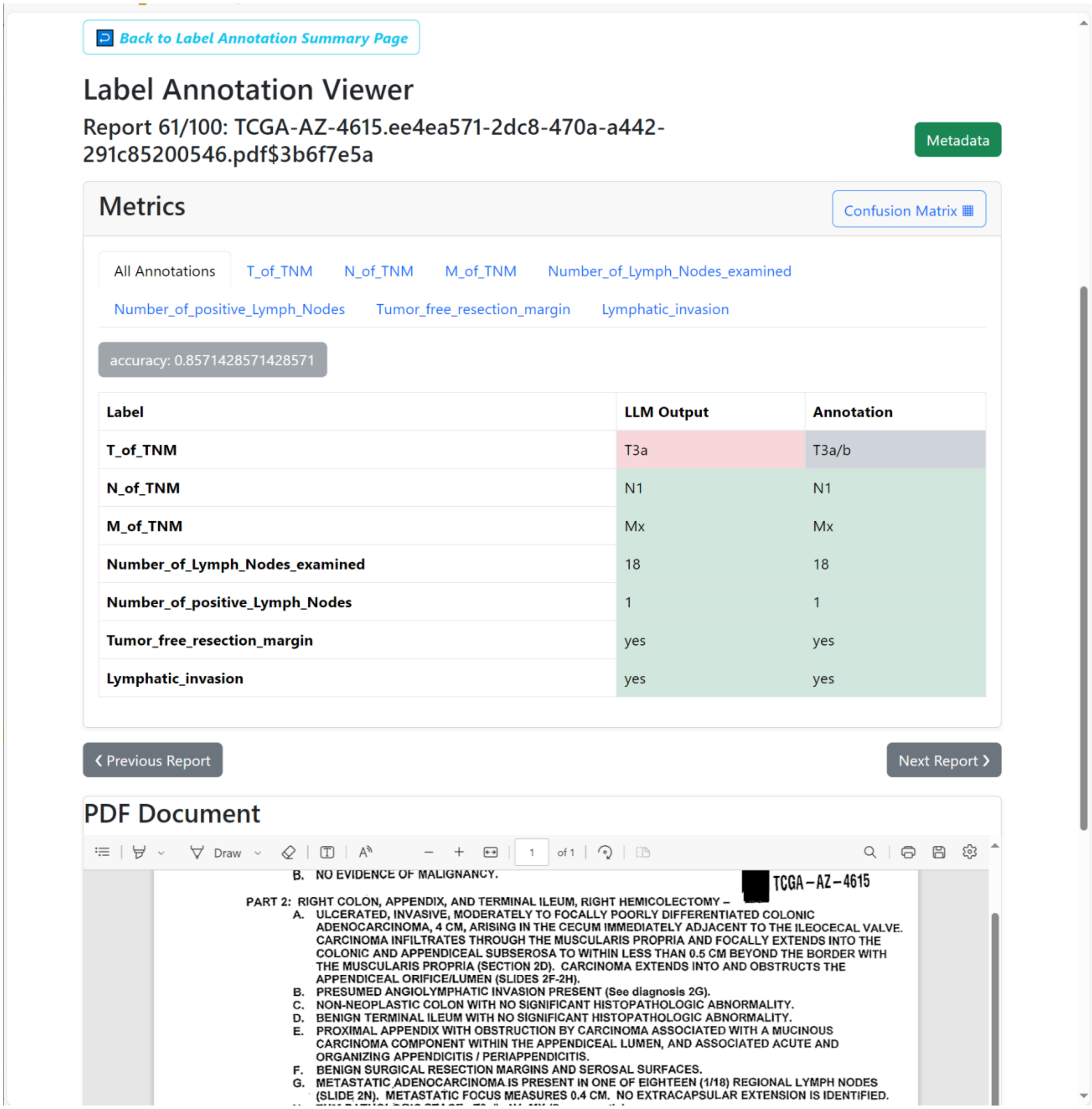


**Supplementary Figure 16 - Annotation is more detailed than LLM.**

The annotation contained a more detailed description of the T-stage than the LLM-output and suggests that definite classification to T3a or T3b was not possible. The LLM output was T3a, which is slightly different but not entirely incorrect when compared to the ground truth.

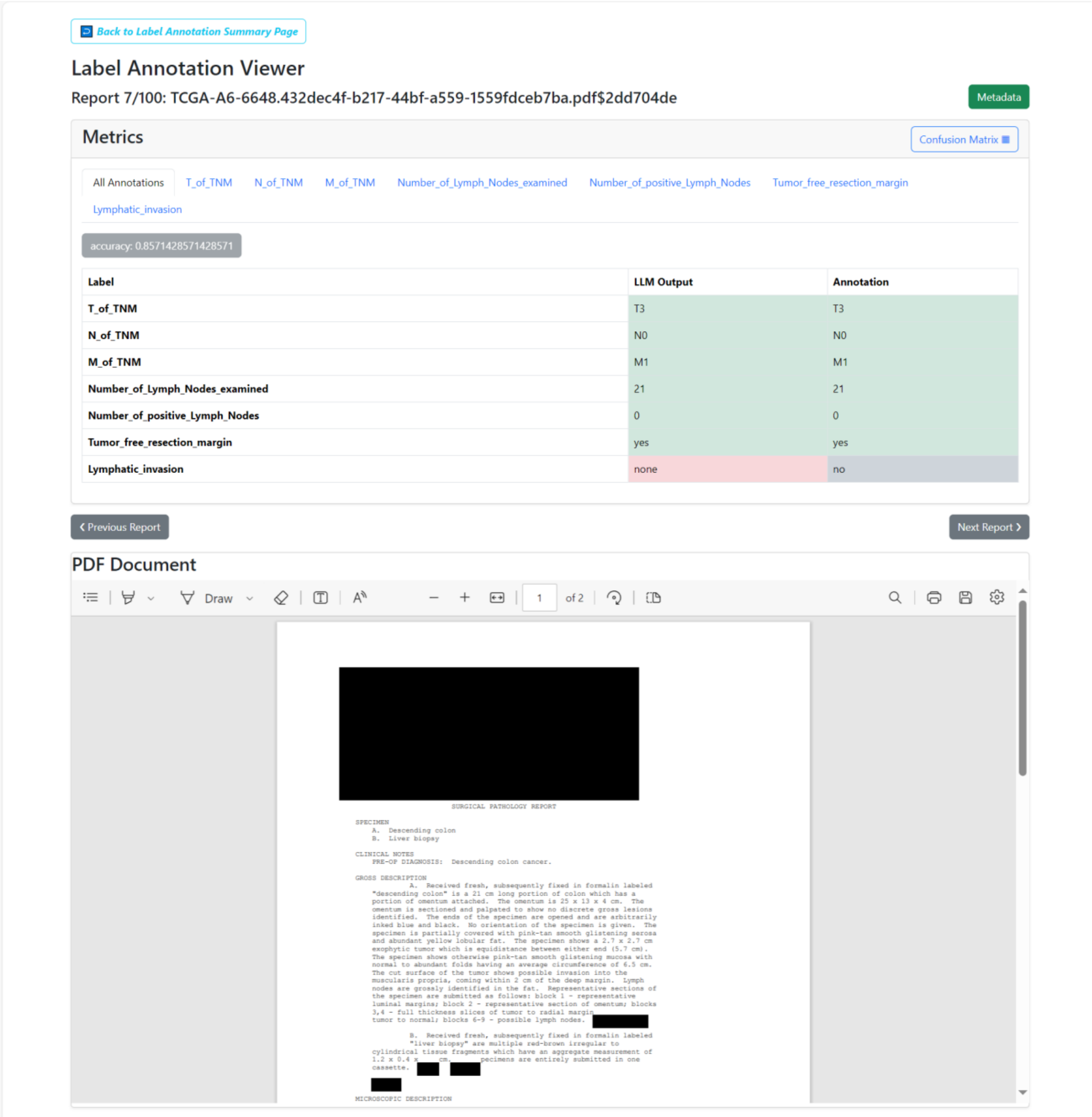


| Label | LLM Output | Annotation |
|---|---|---|
| T_of_TNM | T3 | T3 |
| N_of_TNM | N0 | N0 |
| M_of_TNM | M1 | M1 |
| Number_of_Lymph_Nodes_examined | 21 | 21 |
| Number_of_positive_Lymph_Nodes | 0 | 0 |
| Tumor_free_resection_margin | yes | yes |
| Lymphatic_invasion | none | no |

[redacted]

SURGICAL PATHOLOGY REPORT

SPECIMEN
A. Descending colon
B. Liver biopsy

CLINICAL NOTES
PRE-OP DIAGNOSIS: Descending colon cancer.

GROSS DESCRIPTION
A. Received fresh, subsequently fixed in formalin labeled "descending colon" is a 21 cm long portion of colon which has a portion of omentum attached. The omentum is 25 x 13 x 4 cm. The omentum is sectioned and palpated to show no discrete gross lesions identified. The ends of the specimen are opened and are arbitrarily inked blue and black. No orientation of the specimen is given. The specimen is partially covered with pink-tan smooth glistening serosa and abundant yellow lobular fat. The specimen shows a 2.7 x 2.7 cm exophytic tumor which is equidistance between either end (5.7 cm). The specimen shows otherwise pink-tan smooth glistening mucosa with normal to abundant folds having an average circumference of 6.5 cm. The cut surface of the tumor shows possible invasion into the muscularis propria, coming within 2 cm of the deep margin. Lymph nodes are grossly identified in the fat. Representative sections of the specimen are submitted as follows: block 1 - representative luminal margins; block 2 - representative section of omentum; blocks 3,4 - full thickness slices of tumor to radial margin tumor to normal; blocks 6-9 - possible lymph nodes. [redacted]

B. Received fresh, subsequently fixed in formalin labeled "liver biopsy" are multiple red-brown irregular to cylindrical tissue fragments which have an aggregate measurement of 1.2 x 0.4 x cm. pecimens are entirely submitted in one cassette. [redacted] [redacted]

[redacted]

MICROSCOPIC DESCRIPTION

**Supplementary Figure 17 - Output classes lack sufficient distinguishability.**

In this example, the presence of lymphatic invasion needs to be extracted from TCGA pathology reports of colorectal cancer specimens. Three answer options are given to the LLM: “yes”, ”no”, and “none”, if there is no information at all about lymphatic invasion. These three classes lack sufficient distinguishability in natural language, as “no” and “none” are semantically too close. Improvement can be achieved by defining classes that are more distinguishable in natural language, such as “yes”, “no”, “not mentioned”.

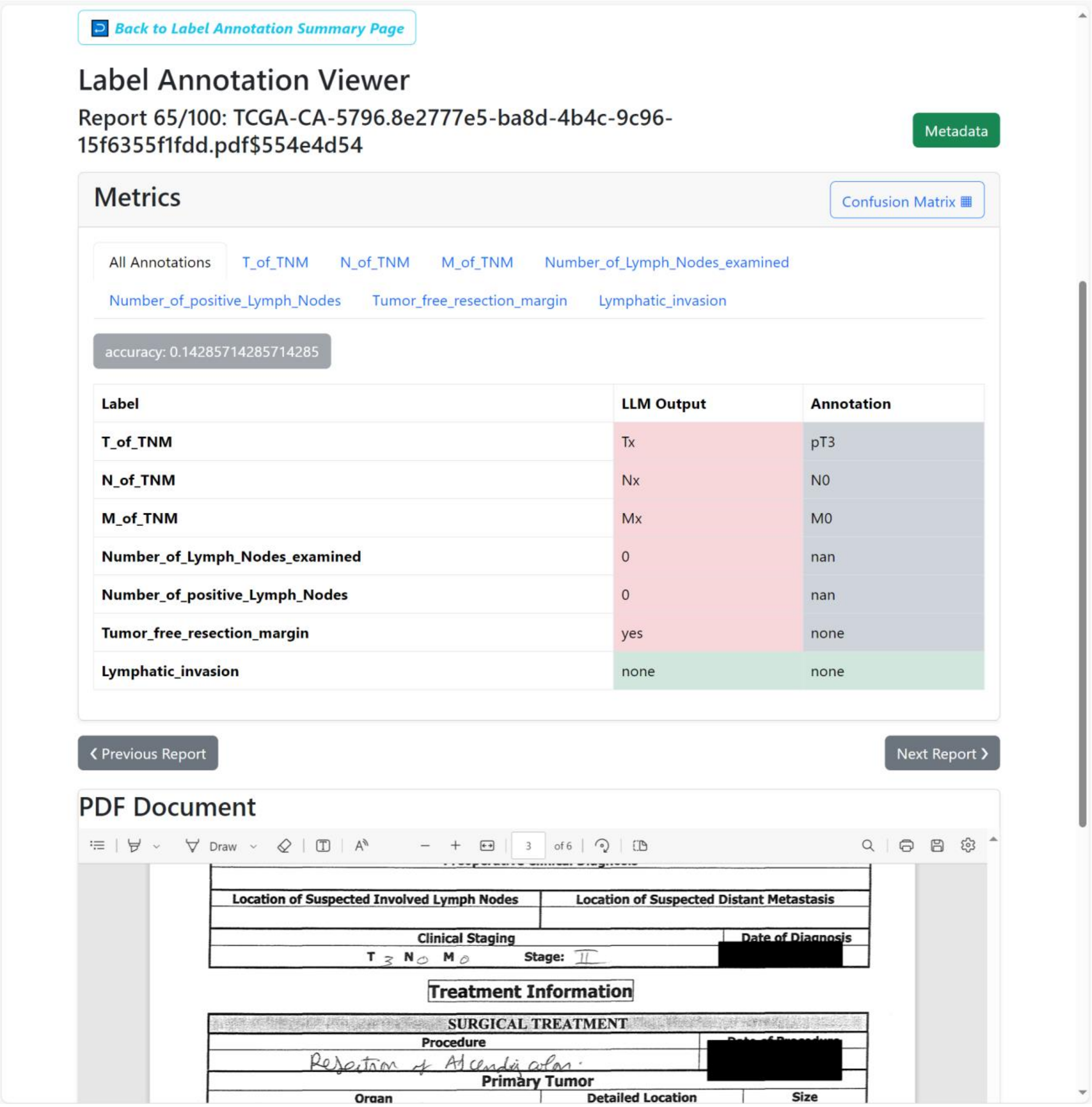


**Supplementary Figure 18 - Failure in information extraction due to bad input data quality.**

When documents contain a high proportion of handwritten information and PDFs are of very poor quality, text extraction may fail due to data preprocessing. This failure can be addressed by choosing an alternative OCR method for preprocessing. The "Surya" OCR method generally outperforms "Tesseract," and trOCR, paddleOCR and visionLLM models such as Phi are superior in detecting handwritten text.

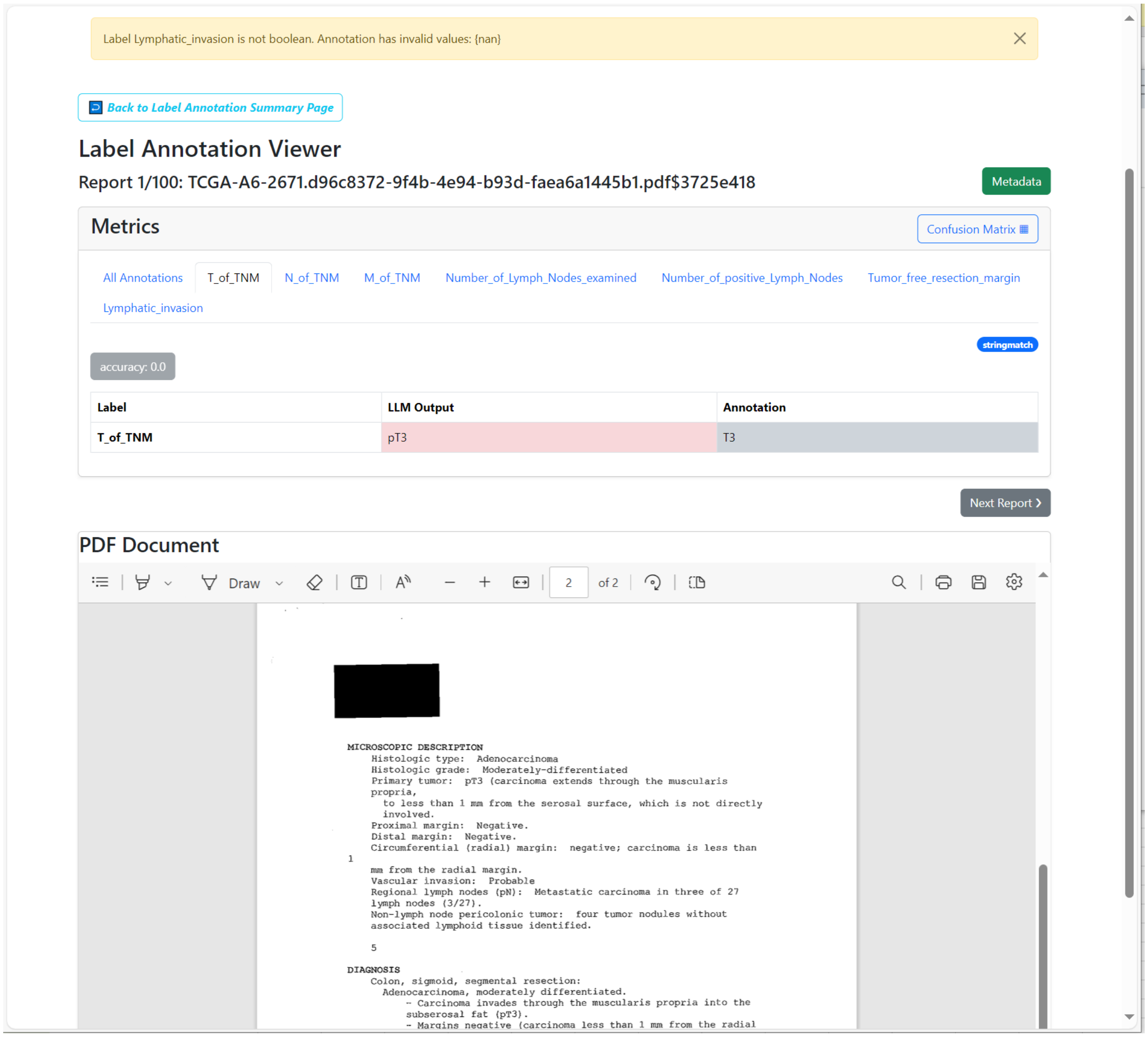


**Supplementary Figure 19 - Accurate LLM output with minor character discrepancies compared to the annotation.**

The report states a “pT3” tumor stage, as the T-stage was defined through histopathological examination of the specimen. The annotation confirms that T3 is correct, but the “p” was not relevant and therefore omitted by the ground truth rater. This discrepancy can be solved via prompt engineering: The desired output needs to be specified as detailed as possible within the prompt and grammar.

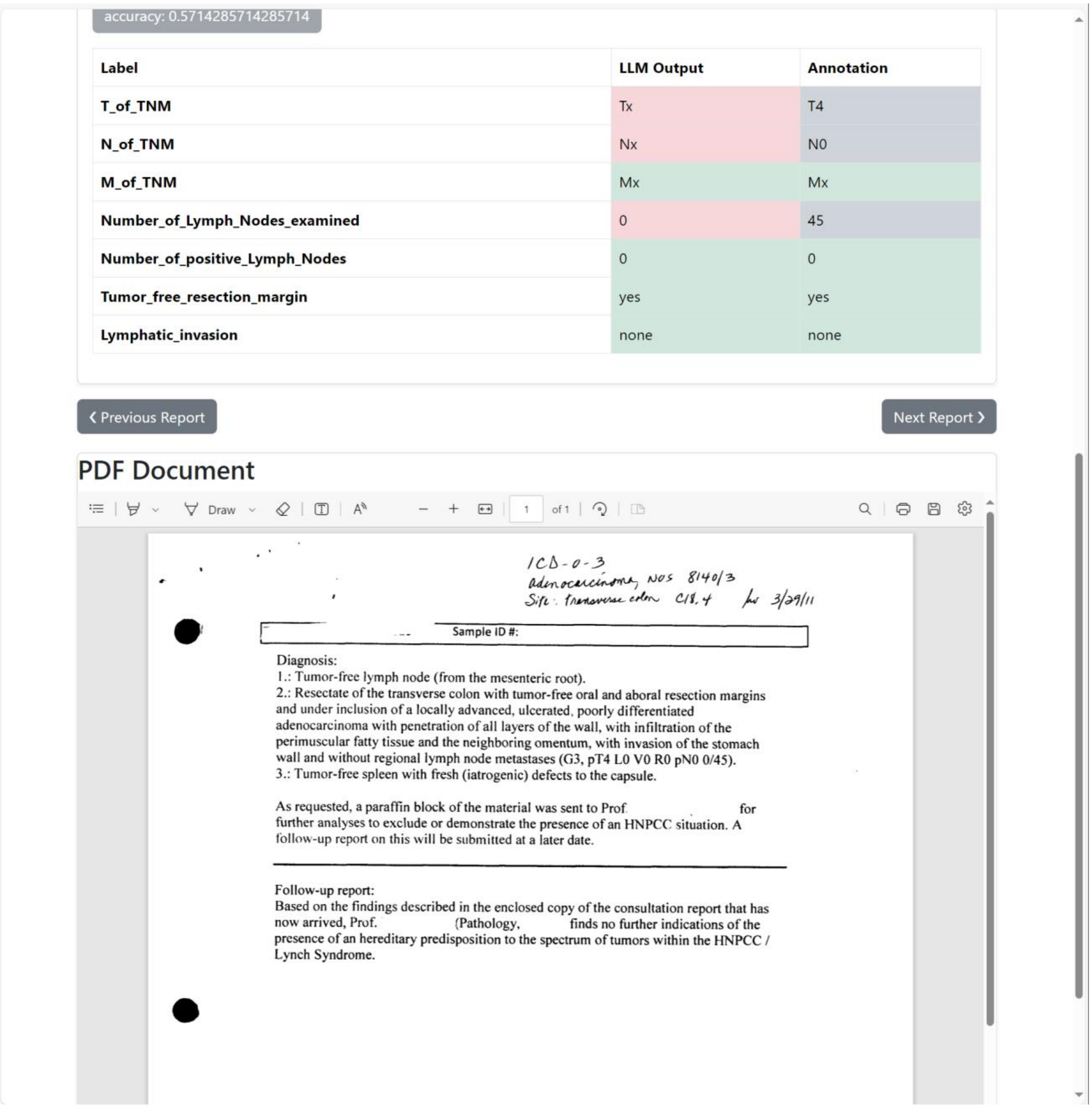

accuracy: 0.5714285714285714

| Label | LLM Output | Annotation |
|---|---|---|
| T_of_TNM | Tx | T4 |
| N_of_TNM | Nx | N0 |
| M_of_TNM | Mx | Mx |
| Number_of_Lymph_Nodes_examined | 0 | 45 |
| Number_of_positive_Lymph_Nodes | 0 | 0 |
| Tumor_free_resection_margin | yes | yes |
| Lymphatic_invasion | none | none |

‹ Previous Report

Next Report ›

PDF Document

1 of 1

ICD-0-3
Adenocarcinoma, NOS 8140/3
Site: transverse colon C18.4 3/29/11

Sample ID #:

Diagnosis:
1.: Tumor-free lymph node (from the mesenteric root).
2.: Resectate of the transverse colon with tumor-free oral and aboral resection margins and under inclusion of a locally advanced, ulcerated, poorly differentiated adenocarcinoma with penetration of all layers of the wall, with infiltration of the perimuscular fatty tissue and the neighboring omentum, with invasion of the stomach wall and without regional lymph node metastases (G3, pT4 L0 V0 R0 pN0 0/45).
3.: Tumor-free spleen with fresh (iatrogenic) defects to the capsule.

As requested, a paraffin block of the material was sent to Prof. for further analyses to exclude or demonstrate the presence of an HNPCC situation. A follow-up report on this will be submitted at a later date.

Follow-up report:
Based on the findings described in the enclosed copy of the consultation report that has now arrived, Prof. (Pathology, finds no further indications of the presence of an hereditary predisposition to the spectrum of tumors within the HNPCC / Lynch Syndrome.

**Supplementary Figure 20 - Information is present but could not be detected by the LLM.**

In this example, the whole tumor formula was given in the text as “G3, pT4 L0 V0 R0 pN0 0/45”. However, this information could not be extracted by the LLM. This could be solved by adding a more detailed explanation of how the tumor formula looks like and few-shot examples in the prompt.

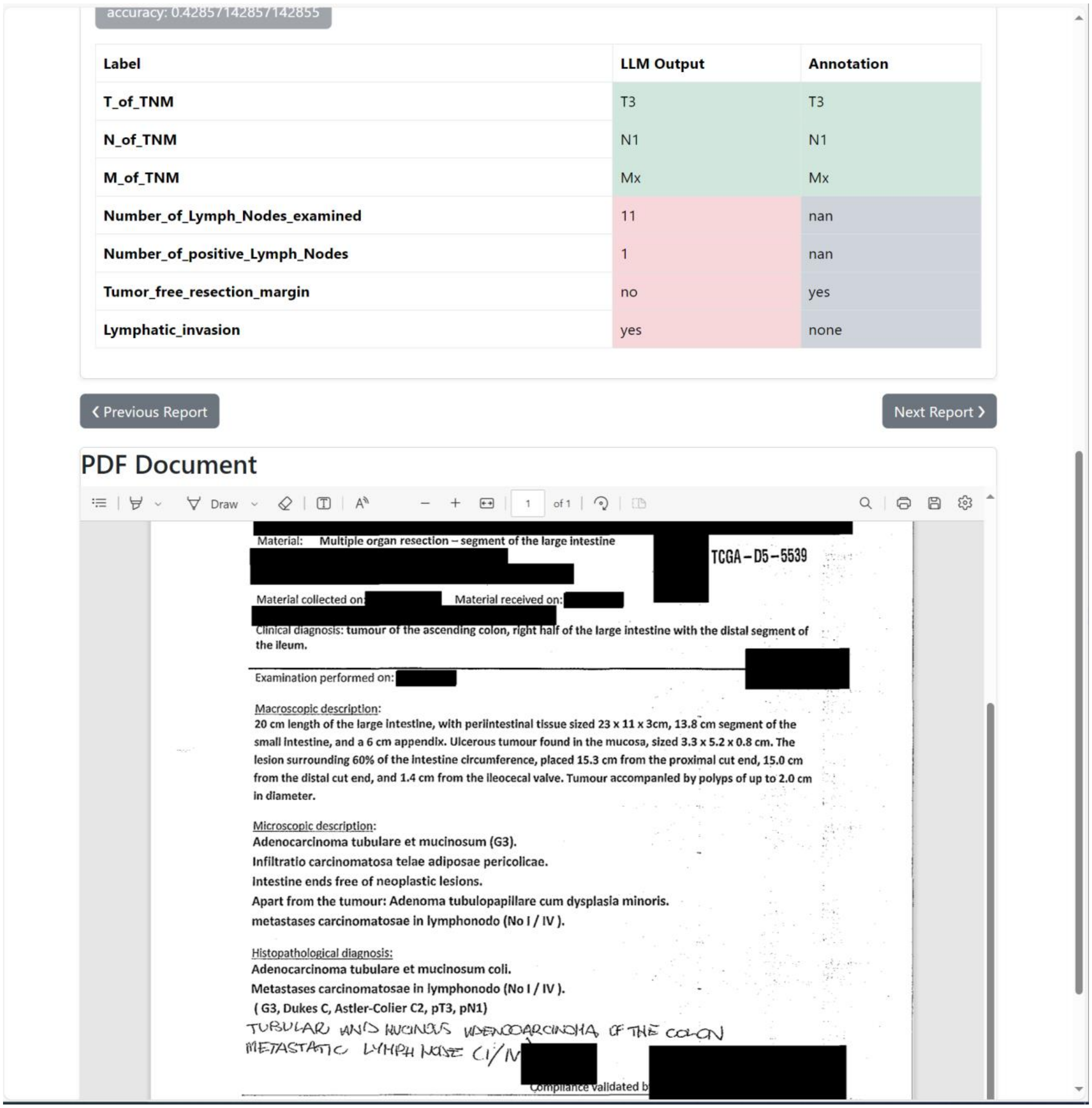

accuracy: 0.42857142857142855

| Label | LLM Output | Annotation |
|---|---|---|
| T_of_TNM | T3 | T3 |
| N_of_TNM | N1 | N1 |
| M_of_TNM | Mx | Mx |
| Number_of_Lymph_Nodes_examined | 11 | nan |
| Number_of_positive_Lymph_Nodes | 1 | nan |
| Tumor_free_resection_margin | no | yes |
| Lymphatic_invasion | yes | none |

‹ Previous Report    Next Report ›

PDF Document

Draw    1 of 1

[redacted]

Material: Multiple organ resection – segment of the large intestine [redacted] TCGA – D5 – 5539

[redacted]

Material collected on: [redacted] Material received on: [redacted]

[redacted]

Clinical diagnosis: tumour of the ascending colon, right half of the large intestine with the distal segment of the ileum.

[redacted]

Examination performed on: [redacted]

Macroscopic description:
20 cm length of the large intestine, with periintestinal tissue sized 23 x 11 x 3cm, 13.8 cm segment of the small intestine, and a 6 cm appendix. Ulcerous tumour found in the mucosa, sized 3.3 x 5.2 x 0.8 cm. The lesion surrounding 60% of the intestine circumference, placed 15.3 cm from the proximal cut end, 15.0 cm from the distal cut end, and 1.4 cm from the ileocecal valve. Tumour accompanied by polyps of up to 2.0 cm in diameter.

Microscopic description:
Adenocarcinoma tubulare et mucinosum (G3).
Infiltratio carcinomatosa telae adiposae pericolicae.
Intestine ends free of neoplastic lesions.
Apart from the tumour: Adenoma tubulopapillare cum dysplasia minoris.
metastases carcinomatosae in lymphonodo (No I / IV ).

Histopathological diagnosis:
Adenocarcinoma tubulare et mucinosum coli.
Metastases carcinomatosae in lymphonodo (No I / IV ).
( G3, Dukes C, Astler-Colier C2, pT3, pN1)

TUBULAR AND MUCINOUS ADENOCARCINOMA OF THE COLON
METASTATIC LYMPH NODE (1/IV [redacted]

Compliance validated b [redacted]

**Supplementary Figure 21 - LLM detects more than human rater.**

In this case, the report mentions “Metastases carcinomatosae in lymphonodo (No I/IV)." This information is also noted in handwriting as “metastatic lymph node (I/IV)." The LLM correctly identifies that one lymph node is positive out of the examined lymph nodes. However, the Roman numeral IV was incorrectly extracted as 11 lymph nodes were examined. This error stems from OCR, where the Roman numeral IV was misinterpreted as II.

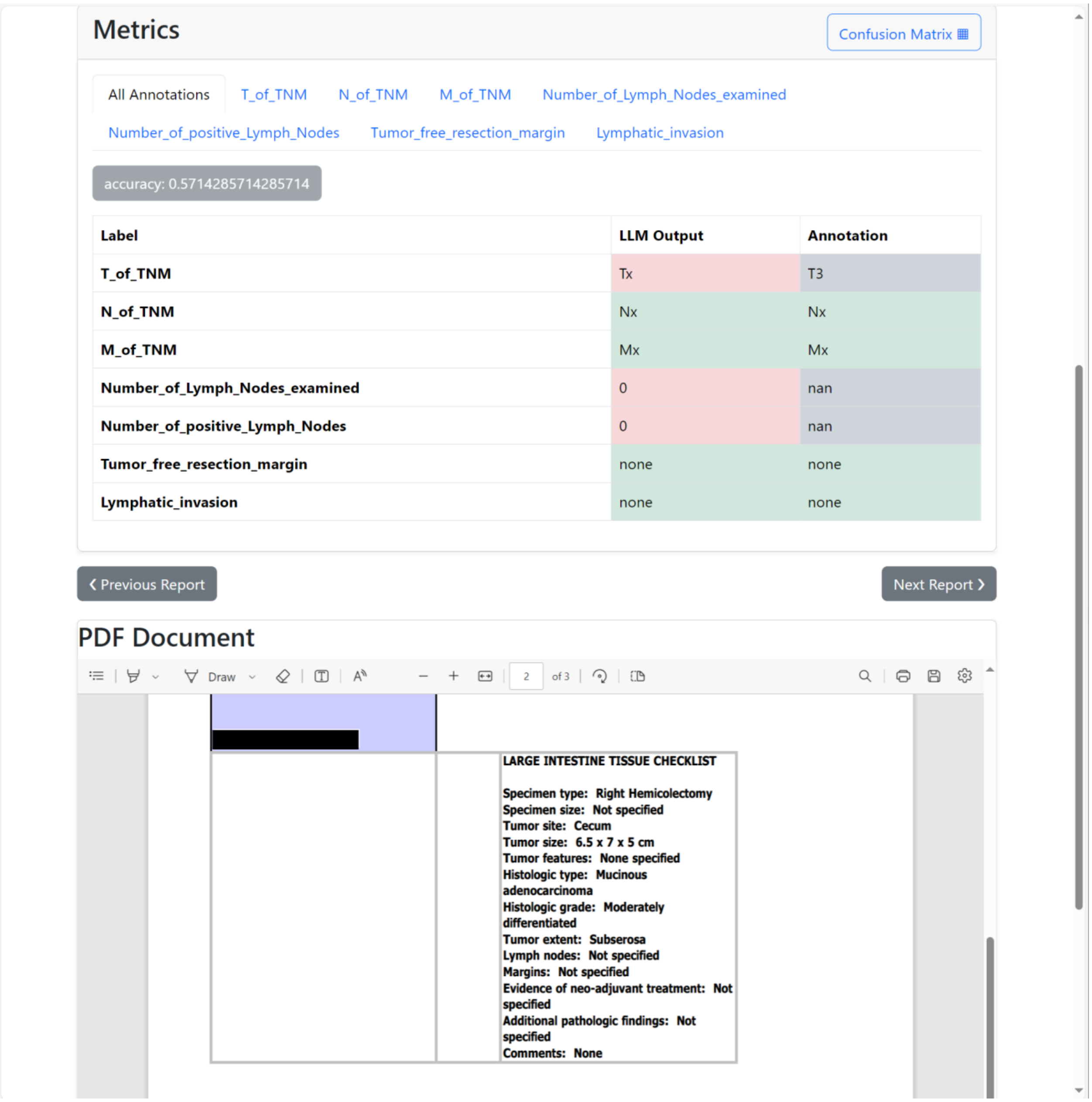


**Supplementary Figure 22 - LLM lacks implicit knowledge.**

The T-stage describes the size and extent of the tumor. The report mentions that tumor extent reaches the subserosa, which corresponds to T3. This was not detected by the LLM and can be solved through more detailed information in the prompt and few-shot examples.

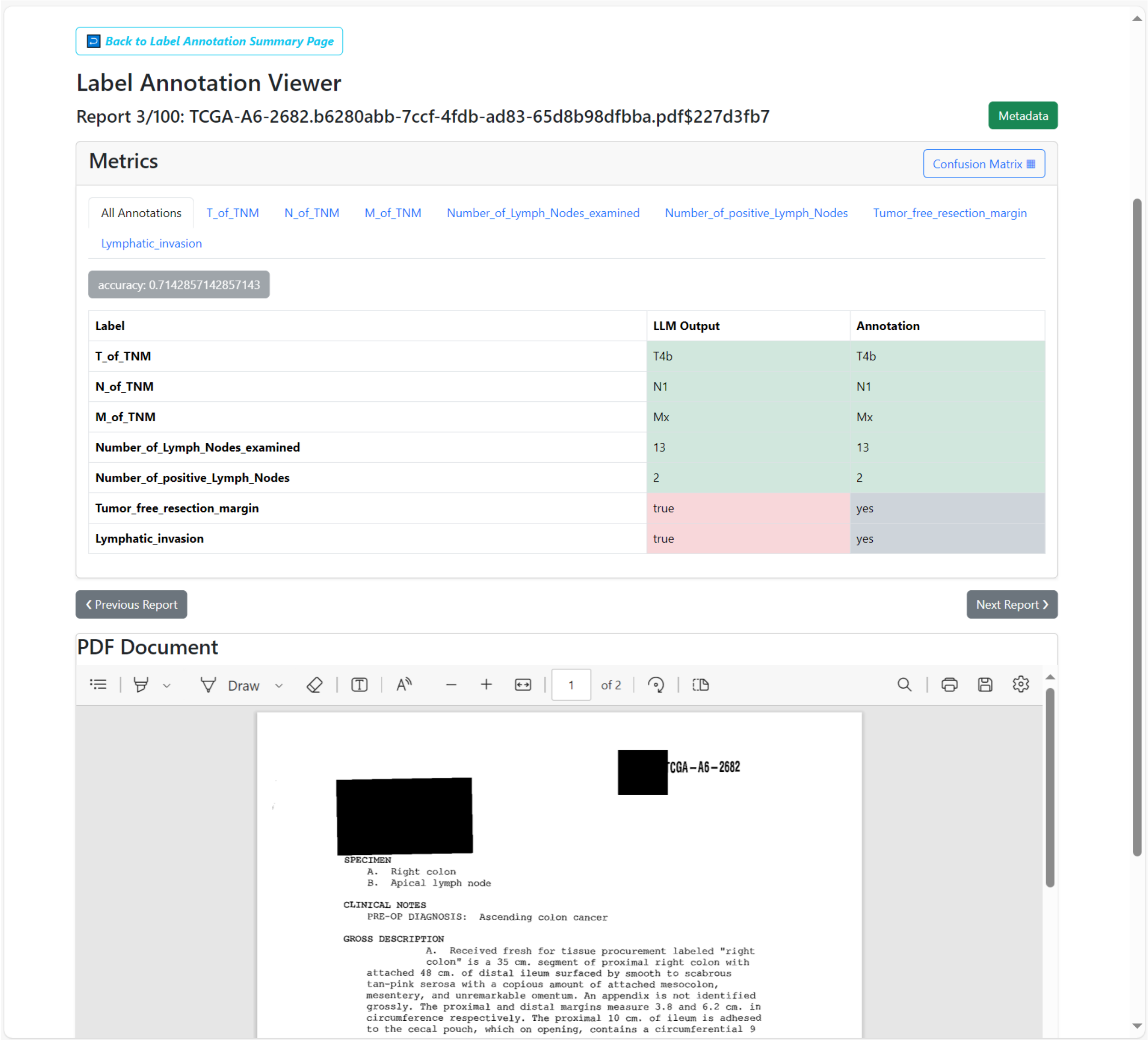


**Supplementary Figure 23 - Mismatch in output categories.**

The output categories must be defined exactly as they appear in the annotated ground truth table. Otherwise, the LLM output might be correct ("true"), but if the ground truth says "yes," it will be considered a mismatch despite having the same meaning.
